# Genome-wide association studies identify shared mechanisms between hypertension and type 2 diabetes independent of adiposity

**DOI:** 10.1101/2025.10.03.25337275

**Authors:** Bethany Voller, Ruby M Woodward, Qingze Gu, Olivia Murrin, João Delgado, Concepción Violán, Sara Khalid, Deniz Türkmen, Chris Fox, Sarah E Lamb, Mary Mancini, Leon Farmer, Kate Boddy, Frank Dudbridge, Jack Bowden, Timothy M Frayling, Jane AH Masoli, Luke C Pilling, the GEMINI Consortium

**Author notes:** joint senior authors (Timothy M Frayling, Jane AH Masoli, Luke C Pilling). **Corresponding author**: Luke Pilling. AGE group, University of Exeter, College House, Magdalen road, Exeter, EX1 2LU, UK.

## Abstract

**Background:** Hypertension and type 2 diabetes (T2D) are two of the most frequently co-occurring long-term conditions, but their shared mechanisms are not fully understood, often being attributed to adiposity pathways. Here, we aimed to identify shared genetic mechanisms independent of adiposity.

**Methods:** We performed genome-wide association study meta-analyses of T2D and, separately, hypertension. We investigated the bidirectional causal relationship using Mendelian randomisation and quantified genetic correlation before and after accounting for common modifiable risk factors. We then applied a Bayesian GWAS approach to re-estimate SNP-disease effects after accounting for the causal genetic effects of adiposity-related traits. Colocalisation analysis identified shared causal genetic variants, and we investigated the biological pathways involved.

**Results:** We observed a bidirectional causal relationship, and substantial genetic correlation between the two traits (rg = 0.48, 95%CI 0.45-0.52), which persisted after accounting for the genetic contributions of BMI, waist-hip ratio (WHR), and triglycerides (rg = 0.29, 95%CI 0.24-0.34). This indicated shared mechanisms beyond those captured by standard measures of adiposity. We found 98 genetic loci containing variants significantly associated with both hypertension and T2D; colocalisation analysis identified 37 that contained specific shared causal variants. Of these, eight remained statistically significant after adjusting for genetic measures of adiposity, and four were identified only after removing the causal effect of adiposity measures. Shared variants include an allele within *PCSK7* associated with risk of both T2D and hypertension and with circulating PCSK7 protein levels, as well as a variant in the 3′ untranslated region of *ZNF101,* within the *TM6SF2* locus, likely reflecting regulatory variation affecting hepatic lipid metabolism and cardiometabolic traits.

**Conclusions:** We identify shared causal mechanisms between hypertension and T2D independent of several measures of adiposity. These findings, particularly PCSK7, could uncover novel interventions or opportunities for prevention of this co-occurring condition pair.

## Introduction

Hypertension and type-2 diabetes (T2D) are two common long-term conditions, that frequently co-exist. Our recent study of over 3 million people showed that individuals with either hypertension or T2D are more than 3 times as likely to have the other condition (Odds Ratio 3.23, 95% Confidence Intervals [95%CI] 3.20-3.25) [1]. Having multiple medical conditions impacts people’s quality of life and increases risk of further adverse health outcomes [2]. Current clinical guidelines and health service delivery focus mostly on treatment and prevention of single diseases rather than conditions co-existing in an individual, contributing to increased rates of polypharmacy, adverse drug events and healthcare use by people with multiple long-term conditions (MLTC) [3,4]. The risk of developing either hypertension or T2D increases with age, health inequalities, and environmental factors including unhealthy diet, alcohol and tobacco consumption, and physical inactivity [5–8]. Obesity, as a marker of excess adiposity, is a key modifiable risk factor for both conditions [9,10]. We previously demonstrated the role of obesity as a driver of MLTC [11].

Despite observational evidence of co-occurrence and high predictive values for hyperglycaemia on hypertension and systolic blood pressure on T2D risk, a previous study using genetic methods only supported the causal effect of T2D on hypertension, but not in the opposite direction [12]. This implies that targeting hyperglycaemia could reduce hypertension risk, but that reducing blood pressure may not reduce T2D risk. The conditions have substantial genetic overlap (genetic correlation r_g_= 48%, 95%CIs 0.45 to 0.52), suggesting shared causal mechanisms, although genetic correlations could also be the result of one condition causing the other [1]. A recent analysis used a genetic approach to identify specific genetic variants that were potentially causal for both type 2 diabetes and coronary artery disease, implicating immune regulation and glucose metabolism pathways—demonstrating the potential of a genetic approach in increasing understanding of shared disease mechanisms [13]. Another recent large study used partitioned polygenic scores to identify clusters of variants associated with measured blood pressure and diagnosis of T2D [14], but no study has specifically identified shared causal variants between diagnosed hypertension and T2D, including those separate to known adiposity-driven pathways.

In this study, we investigate the shared genetic architecture underlying the co-occurrence of hypertension and T2D. We know that the two conditions are genetically correlated [1]: here we assess whether there are bidirectional causal relationships using Mendelian randomisation, a statistical causal inference method that (under specific assumptions) uses genetic variants associated with a trait of interest as instrumental variables to investigate whether the trait has a causal effect on a second trait. We then explore the genetic contribution of common, modifiable risk factors to the genetic relationship, and uncover shared genetic mechanisms acting through pathways distinct to known risk factors. Understanding the mechanistic overlap between this frequently co-occurring condition pair could uncover pathways for prevention and support treatment optimisation, including drug discovery.

## Methods

### Data and sources

The GEMINI (Genetic Evaluation of Multimorbidity towards INdividualisation of Interventions) collaborative investigated novel links between long-term conditions to inform interventions and improve patient outcomes. The first GEMINI study ascertained 72 common, chronic long-term conditions with significant heritability [1]. See GitHub for curated diagnostic code lists (https://github.com/GEMINI-multimorbidity) and website for interactive results (https://gemini-multimorbidity.shinyapps.io/atlas). Observational analysis was based on Longitudinal cohort study using data from the Clinical Practice Research Datalink Aurum (CPRD-Aurum); a UK primary care database comprising anonymised electronic health records (EHRs) from over 17 million active National Health Service (NHS) patients across more than 700 primary care practices [15].

To maximise genetic sample sizes when analysing each condition, we meta-analysed genome-wide association study (GWAS) data from three sources: two large cohort studies UK Biobank (N=502,000), FinnGen (release 9; N= 377,000) [16] and condition-specific consortium data when available [1]. Analyses only included participants genetically similar to the 1000 Genomes European (EUR) subset, to enable using Linkage Disequilibrium Score Regression (LDSC, described below) [17].

For hypertension, we meta-analysed GWAS from UK Biobank (N diagnosed=158,746, N controls=291,451) and FinnGen (N diagnosed=111,581, N controls=280,842). No public GWAS consortia meta-analysis was available at the time. For T2D, we meta-analysed a published GWAS consortium meta-analysis (EUR subset N cases=80,154, N controls=853,816) [18] with data from FinnGen (N diagnosed=57,698, N controls=334,725) (the consortium GWAS already included UK Biobank). This gave effective sample sizes for meta-analysis of 725,376 for hypertension and 446,142 for T2D. There is therefore substantial sample overlap (>50%) as UK Biobank and FinnGen are included in both GWAS. GEMINI GWAS summary statistics are available (https://doi.org/10.5281/zenodo.14284046).

We used summary statistics of recently published GWAS from leading consortia for 10 risk factors: BMI, waist-hip ratio, cholesterol (LDL, HDL, triglycerides), educational attainment, smoking status (ever smoked, and separately smoking intensity in smokers), alcohol consumption, and physical activity (see Supplementary Information for details).

### The genetic relationship between hypertension and type 2 diabetes

Linkage Disequilibrium-score regression (LDSC) was used to estimate the genetic correlation between hypertension and T2D. LD-scores for LDSC were previously estimated from the 1000 Genomes European reference panel [17]. Genetic correlation is an estimate of the correlation of effects across all causal single nucleotide polymorphisms (SNPs) in the genome between two traits. It is a measure of pleiotropy – when a genetic locus affects more than one trait. Genetic correlation can reflect different scenarios: there could be a causal relationship between the two traits with one causing the other, or there could be a common risk factor or biological pathway that causes both traits, with or without a causal relationship between them [19].

To further investigate the relationship between hypertension and T2D, we used Mendelian randomisation (MR) to estimate bidirectional causal effects. MR is a statistical method that allows us to estimate the causal effect of an exposure on an outcome, using genetic variants as instrumental variables (IVs), under certain assumptions (Supplementary Figure 1) [20]. MR studies can make stronger inferences on causality than observational studies; observational studies are susceptible to confounding, reverse causation, and other biases, limiting the validity of their findings. Genetic variants are fixed at conception and randomly inherited, making them less prone to these problems [21]. We performed two-sample MR, using the TwoSampleMR R-package (version 0.6.6) [22], focusing on the inverse-variance weighted (IVW) estimate for the primary analysis.

To select the genetic variants as instrumental variables for the exposure, we identified the genome-wide significant variants (p < 5*10^-8^), and estimated independent genetic loci based on a distance of 500kb using the function ‘get_loci’ from the R-package ‘gwasRtools’ (version 0.1.7, https://github.com/lcpilling/gwasRtools). The SNP with the smallest p-value in each locus was labelled as the lead SNP; these were filtered to those present in the outcome GWAS summary statistics and selected as IVs. Both the exposure and outcome GWAS were of individuals of European ancestry, minimising the risk of population stratification and differences in LD structure. Both included adults of similar age and sex distributions, reducing the likelihood of context-dependent genetic effects.

For a binary exposure, such as hypertension or T2D, the IVs used in an MR analysis capture the liability to the exposure, rather than variation in the exposure [23,24]. It has been recommended that MR analyses using a binary exposure should be used as a test of the causal null hypothesis, rather than an attempt to calculate a causal estimate [24,25]. For binary disease outcomes, the causal estimates are on the log odds ratio scale, representing the change in the log odds of the outcome per unit increase in the log odds of the exposure. To improve interpretability, we converted estimates so they represent the change in the odds of the outcome per doubling in odds of the exposure; this was obtained by multiplying the causal log OR estimates by log_e_2 prior to exponentiating [25].

The validity of findings from MR analyses relies on three IV assumptions being satisfied (IV1: relevance, IV2: independence, and IV3: exclusion-restriction; illustrated in Supplementary Figure 1), and two of these cannot be fully empirically tested (IV2 and IV3). We performed extensive sensitivity analyses to investigate robustness of findings to variations in model assumptions, including effects of pleiotropy (balanced and unbalanced). Relaxing the exclusion-restriction assumption (the IVs are only associated with the outcome through the exposure), we looked for consistency of the IVW estimate with the MR-Egger, weighted median and weighted mode estimates; these are slightly more robust to pleiotropy [26–28]. We included the recently proposed MR-GRIP method, which is invariant to allele coding, unlike MR-Egger [29]. We used Cochrane’s Q-statistic, a measure of heterogeneity, to test for global pleiotropy [30]. Where heterogeneity was detected (Q > number of IVs –1), we identified any outlying variants using Radial MR. Here, each variant’s contribution to Cochrane’s Q-statistic is quantified, and outliers are detected using a significance threshold of 0.05 [31]. The exposure-outcome association was re-estimated following removal of any outliers. Steiger filtering was used to assess the validity of the IVs by identifying and removing variants that had a stronger association with the outcome than the exposure [32]. This study complies with the STROBE-MR (‘Strengthening the Reporting of Observational Studies in Epidemiology using Mendelian Randomisation’) guidelines for reporting MR [33].

### The contribution of risk factors to shared genetics

To quantify the genetic contribution of common risk factors to the shared genetics of hypertension and T2D, we used our recently published methodology [34], partial LDSC, implemented in an R-package (https://github.com/GEMINI-multimorbidity/partialLDSC). Our method takes GWAS summary statistics and computes partial genetic correlations, defined as the genetic correlation between two conditions, holding the genetic effects of potential confounders constant [34]. This gives an estimation of the genetic correlation of two conditions that does not act through any additional specified traits, as estimated by the genetic data currently available for those traits. In our original paper, the method is used to highlight obesity as a key mechanistic cause for many LTC pairs. Here, we extend partial LDSC to investigate the contribution of 10 common risk factors, both individually and in a multivariate manner, to the shared genetics of hypertension and T2D (R package {partialLDSC} v0.2.0 - see Supplementary Methods for details).

### Removing the causal effects of risk factors using genetics

We examined results from partial LDSC analyses to assess which risk factors had the biggest effects on the shared genetic association. We then used a Bayesian GWAS approach, implemented in the bGWAS R-package (version 1.0.3), to derive direct effect estimates: estimates of the causal effect of each SNP, independent of the causal effects of the identified risk factors, on hypertension and T2D respectively [35]. Variants with a significant direct effect estimate are likely to affect the condition either directly, or via pathways independent of the included risk factors. We selected instruments for each risk factor using a threshold of p < 5*10^-8^. Direct effect estimates from bGWAS analyses were output as GWAS summary statistics.

### Identifying shared causal mechanisms using genetic colocalisation

We identified the variants significantly associated (p < 5*10^-8^) with both hypertension and T2D. We then defined regions of ±250kb around each variant, starting with the variant with the smallest p-value. This was done iteratively until all variants had been assigned a region. We then performed statistical colocalisation using the *coloc.abf* function from the coloc R-package (version 5.2.3) [36], to investigate whether hypertension and T2D shared causal variants. We did this first using the original hypertension and T2D GWAS summary statistics, and then again using the bGWAS summary statistics to see whether any shared causal variants remained after removing the causal effects of common, known risk factors.

Coloc is a Bayesian method and works by calculating the posterior probabilities of five hypotheses for each given genomic region; we focused on hypotheses 3 and 4 (H3 and H4), which are: H3: there are two distinct causal variants in the region, one for each trait, and H4: there is a single causal variant in the region that is common to both traits [36]. We used the default prior probabilities: p_1_ =1 x 10^-4^, p_2_ =1 x 10^-4^, p_12_ =1 x 10^-5^, where p_1_ and p_2_ are the probabilities that a random SNP in the region is causally associated with trait 1 or trait 2 respectively, and p_12_is the probability that a variant is causal for both traits. The default value p_12_ =1 x 10^-5^ is more likely to detect false shared associations than a more conservative choice of p_12_, but may also lead to missing valid shared causal variants with slightly weaker signals [37].

We considered evidence for colocalisation sufficient if the combined posterior probability of H3 and H4 (PPH3 and PPH4, respectively) was high: PPH3+PPH4 ≥ 0.8 – this suggests an association with both traits, and if PPH4/PPH3 > 5 – suggesting sufficient support to consider the colocalising signal ‘convincing’ [38]. For each region with evidence of colocalisation, we extracted the posterior probabilities for each SNP in the region to be causal conditional on H4 being true.

### Follow up of shared signals including proteomics

We carried forward the SNPs with PPH4 > 0.5 as the SNPs most likely to be causal for both conditions. Where identified variants were within 2.5Mb we used GCTA-COJO to determine whether variants are independent (using 5,000 unrelated UK Biobank participants of European genetic ancestry as LD reference data) [39]. We searched each SNP in the European Bioinformatics Institute (EBI) GWAS Catalog (https://www.ebi.ac.uk/gwas/) [40]. We also searched the GWAS Catalog for traits associated with variants in strong linkage-disequilibrium (LD) (R^2^ > 0.9) with each of our identified SNPs. The Open Targets Platform (https://platform.opentargets.org/) was used to investigate genetic associations from the literature, along with the genes linked to the variant [41]. The GTEx Portal (https://www.gtexportal.org/home/) was used to find expression quantitative trait loci (eQTL) evidence for each variant [42]. The UCSC Genome Browser was used to explore the genomic position of each variant in relation to surrounding genes (genome build hg19) [43]. GeneCards was used to look up the implicated genes [44], and current literature was searched for both the genes and variants.

We used proteomic data for 2,923 proteins from the UK Biobank Pharma Proteomics Project (N=48,195 European individuals), profiled using Olink technology, to investigate whether variants were protein quantitative trait loci (pQTLs) at p < 5*10^-5^ [45]. We did this for the SNPs identified in the colocalisation analysis after adjusting for the causal effects of risk factors. We classified *cis*-pQTLs as those residing within 50kb of the gene transcription start site; the rest we defined as *trans*-pQTLs.

We investigated the druggable status of identified proteins using the Druggable Genome database; the ‘druggable status’ of a protein describes the ability of the protein to bind to a typical drug-like molecule with high affinity [46]. We used DrugBank and the drug-gene interaction database to see whether there were approved drugs, or drugs in clinical trials for potentially druggable proteins, and if so, whether they were known to be used for hypertension and/or T2D [47,48].

### Observational data

We conducted observational analysis to link our genetic analysis with real-world health data to inform potential opportunities for prevention in the translational pathway. We included CPRD Aurum patient data on participants registered with a general practice who were alive and aged ≥40 at any time between 01/01/2010 and 01/01/2020. The study-start-date was the earliest date in which the criteria was met. Diagnosis of hypertension and type 2 diabetes (T2D) cases, and comorbidities (chronic kidney disease, obesity) [1], were defined by prevalent Read V2 or SNOMED-CT codes at any time before the study-start-date. Lifestyle factors (smoking status, alcohol consumption, physical activity), and clinical measures (height, weight, body mass index [BMI], systolic and diastolic blood pressure [SBP, DBP], high-density and low-density lipoprotein cholesterol [HDL, LDL]), were retrieved if recorded in the 5 years prior to study-start-date. Index of Multiple Deprivation [IMD] quintile) was based on ONS practice postcode and demographic characteristics (gender, ethnicity), socioeconomic status (region of residence) where extracted from patient record information.

Three mutually exclusive cohorts were defined: (1) hypertension-only (N=2,443,295), (2) T2D-only (N=395,210), and (3) coexisting hypertension and T2D (N=755,706) (see Table S1). For individuals with both conditions, the order of disease onset was determined by comparing the first recorded diagnosis dates, with the interval between diagnoses reported in years.

Binomial proportion tests were used to compare the probability of hypertension-first versus T2D-first diagnoses, and differences in time to diagnoses were assessed using t-tests or Wilcoxon rank-sum tests, as appropriate. Co-occurrence patterns of hypertension and T2D were visualised using an UpSet plot.

### Patient and Public Involvement (PPI)

Two co-authors, LF and MM, are public collaborators with direct experience of living with multiple long-term conditions. As co-investigators they worked in partnership with the whole research team by attending fortnightly research meetings to progress this research. Five online PPI meetings for additional patients and carers with experience of multiple long-term conditions were held during the study life course to inform decision making. These meetings contextualised the importance of the research from a patient perspective, directly informed research decisions on the selection of multiple long-term conditions for analysis, and elicited outcome priorities for patients. Further information about the PPI approach and the impact of these meeting can be found here https://sites.exeter.ac.uk/gemini/get-involved/.

### Ethics statement

Data from the Clinical Practice Research Datalink (CPRD) were obtained under license from the UK Medicines and Healthcare products Regulatory Agency. The data are provided by patients and collected by the NHS as part of their care and support. Approval for CPRD data access was granted by the CPRD Independent Scientific Advisory Committee, protocol number 23_003109. Anonymised routinely collected healthcare data were used for all CPRD analyses; therefore, informed consent was not necessary.

Individual level UK Biobank genetic data was used for investigating genetic variant co-inheritance and Linkage Disequilibrium in colocalisation and conditional analyses. The Northwest Multi-Centre Research Ethics Committee approved the collection and use of UK Biobank data (Research Ethics Committee reference 11/NW/0382). Participants gave informed consent for the use of their data, health records, and biological materials for health-related research purposes. Access to UK Biobank was granted under Application Number 14631.

## Results

### Genome wide association study meta-analyses

We separately meta-analysed GWAS summary statistics for hypertension and T2D [1]. We identified genome-wide significant (p < 5*10^-8^) variants for each condition (N=46,039 and 28,222, respectively). We then used a distance-based approach to define genomic loci, grouping variants ±250kb around the most significant variants, resulting in 754 and 590 loci, respectively. We next detail the analyses undertaken to identify shared signals between the two conditions, before and after accounting for adiposity, which are summarised as a flowchart in Figure 1.

**Figure 1:**
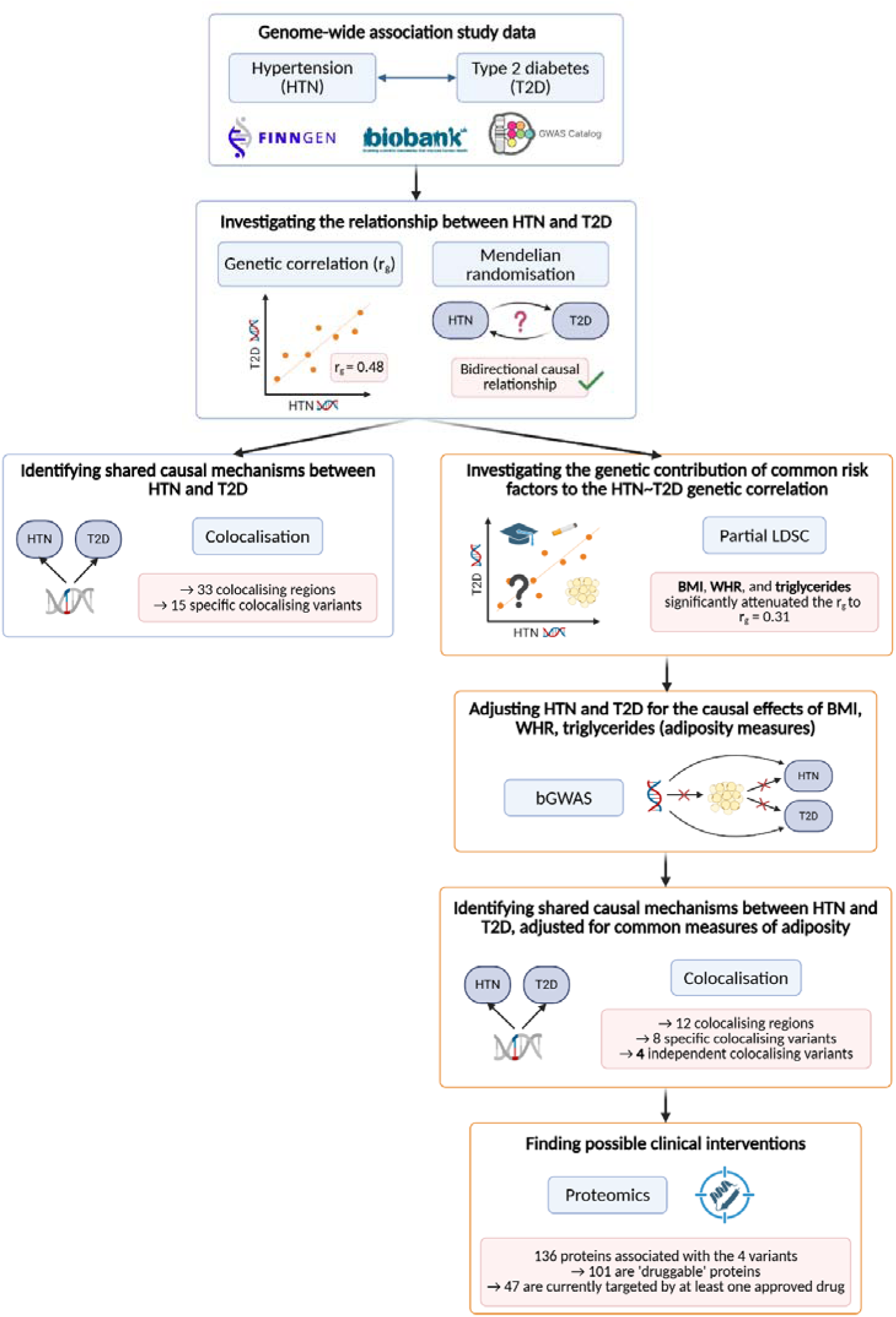
Flowchart of genetic analyses. Flowchart illustrating the steps of the genetic analyses performed, along with key results. Orange boxes indicate analyses investigating the role of shared risk factors on shared mechanisms. Created in BioRender. Voller, B. (2025) https://BioRender.com/3q4njma.

### Genetic relationship between hypertension and T2D

Genetic correlation analyses showed hypertension and T2D were strongly genetically correlated (r_g_ = 0.48, SE = 0.02, p = 2.07*10^-196^), suggesting shared genetic mechanisms. This reflected an estimated 48% positive correlation between the effect sizes of all SNPs across the genome included in the analysis on hypertension and T2D, suggesting shared genetic mechanisms.

### Estimated causal effect of hypertension on T2D

Mendelian randomisation (MR) analyses tested if genetic correlation reflected a causal relationship between hypertension and T2D. MR analysis supports a causal effect of hypertension on T2D: a doubling in genetically instrumented liability to hypertension increased risk of T2D by 1.24x (IVW OR = 1.24; 95% CI: 1.21-1.27; p = 2.04*10^-57^) (Figure 2), though there was substantial evidence of heterogeneity of estimates from the genetic variants used to instrument hypertension. The estimated effect of hypertension on T2D was slightly smaller in sensitivity analyses, using pleiotropy-robust methods, and excluding variants with disproportionately large or small effects on the MR estimates (OR = 1.21; 95% CI: 1.20-1.23; p = 2.23*10^-145^). See Supplementary Table 1 for variants selected as IVs, Supplementary Table 2 for bidirectional MR results and sensitivity analyses, Supplementary Results for additional sensitivity analyses, and Supplementary Figures 2 and 3 for details.

**Figure 2:**
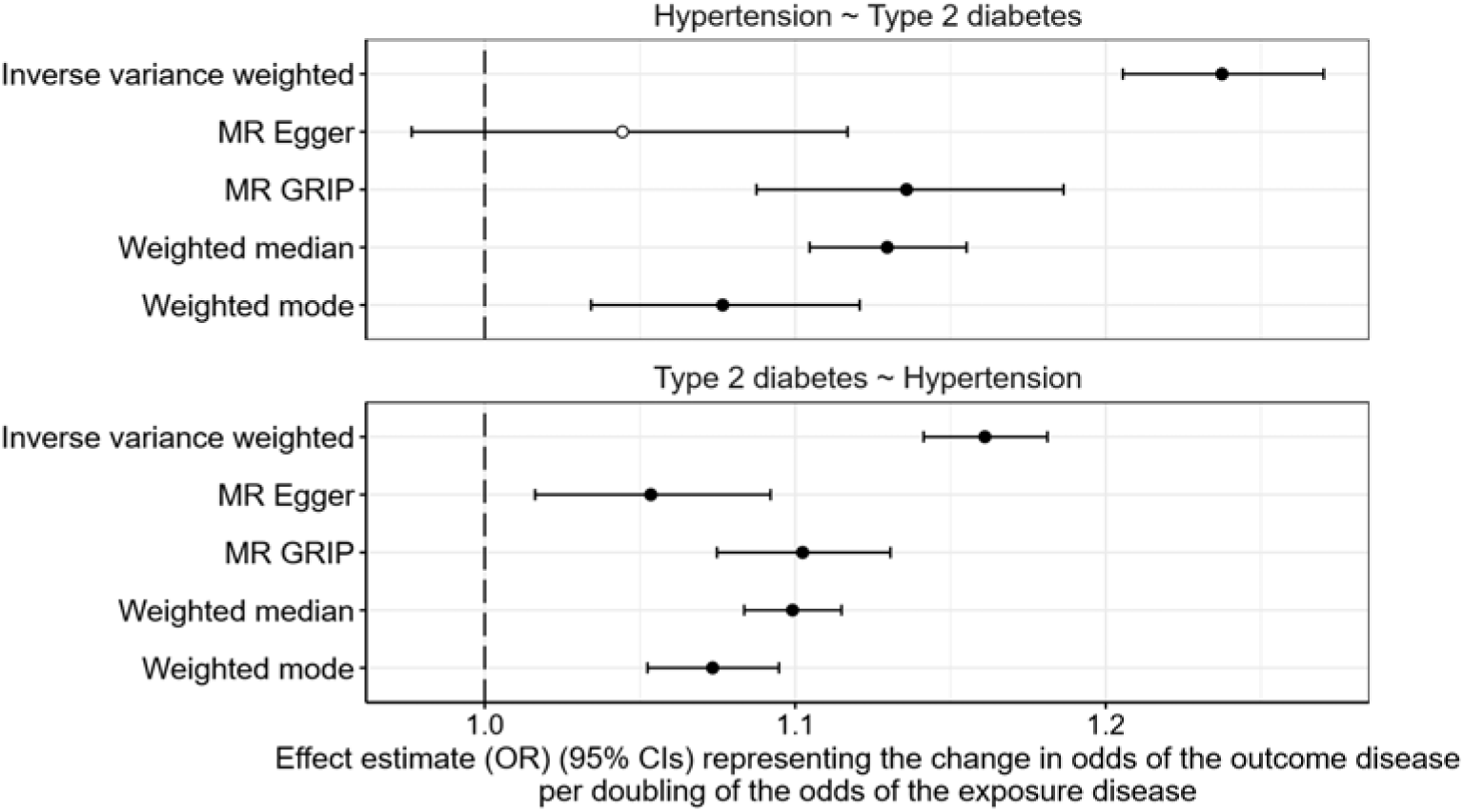
Genetic analysis supports bidirectional causal effects between hypertension and type 2 diabetes. Bidirectional MR estimates for hypertension and T2D. The effect estimates denote the change in odds of the outcome per doubling of the odds of the exposure.

**Figure 3:**
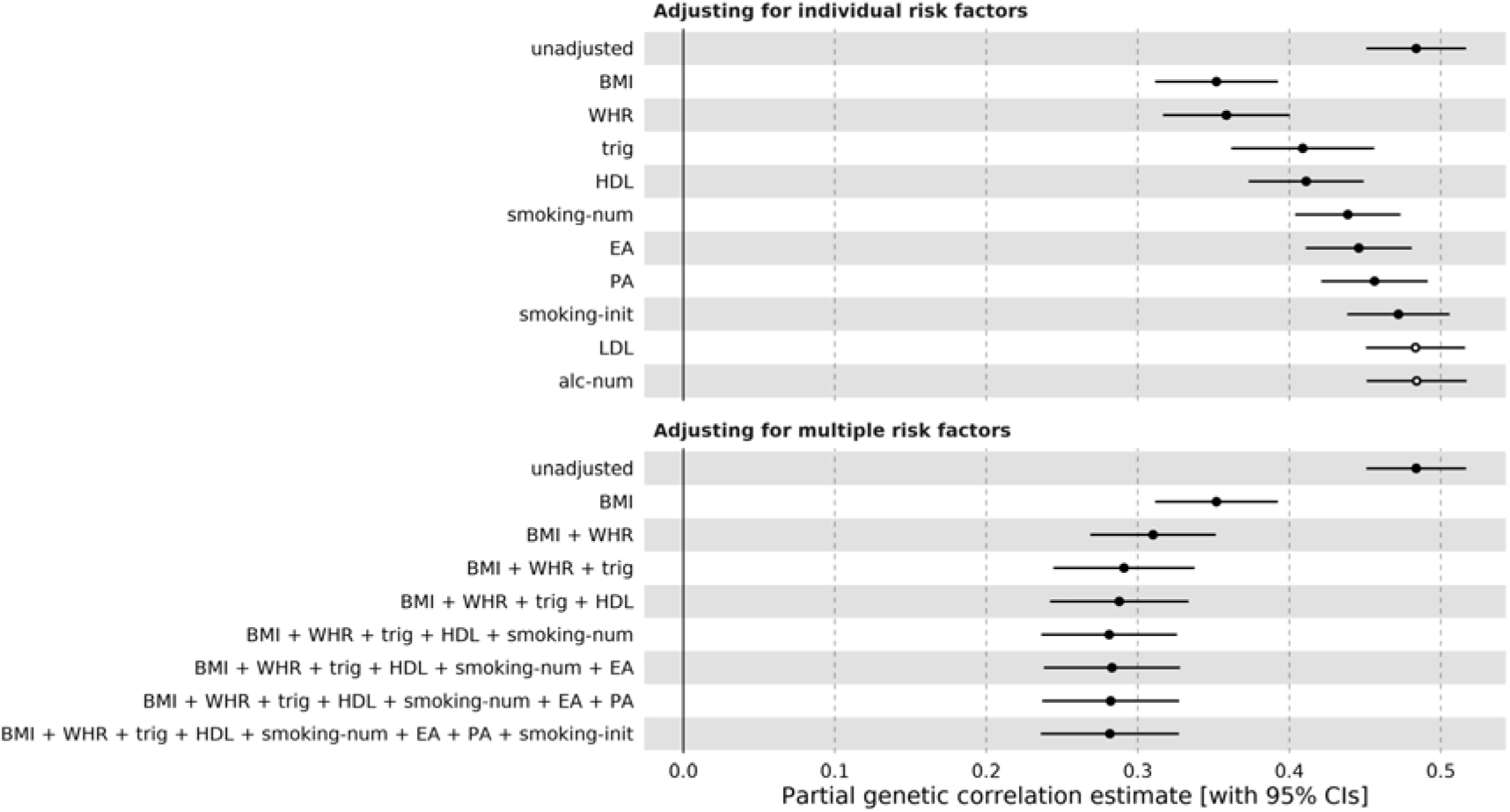
The contribution of modifiable risk factors to the genetic correlation between Hypertension and Type 2 Diabetes. Partial genetic correlation results, showing remaining genetic correlation between hypertension and T2D after adjusting for the genetic of risk factors. The top panel shows the univariable results; each risk factor is individually adjusted for. The bottom panel shows the multivariable results; the risk factors are adjusted for in an additive way, starting with the risk factor with the largest attenuation of the initial hypertension-T2D genetic correlation. A filled circle indicates statistical significance for the difference in genetic correlation. BMI = body mass index; WHR = waist-to-hip ratio; LDL = LDL-cholesterol; HDL = HDL-cholesterol; trig = triglycerides; EA = educational attainment; smoking-init = smoking initiation; smoking-num = smoking (number per day); alc-num = alcohol (drinks per week); PA = physical activity.

### Estimated causal effect of T2D on hypertension

MR analysis also supported a causal effect of T2D on hypertension: IVW analysis showed that a doubling in genetically instrumented odds of T2D increased risk of hypertension by 1.16x (IVW OR = 1.16, 95% CI: 1.14-1.18; p = 2.40*10^-65^) (Figure 2). The association was consistent after removal of outlier variants contributing to heterogeneity, and in other sensitivity analyses (see Supplementary Table 1, Supplementary Table 2, Supplementary Results, and Supplementary Figures 2 and 3).

### Effect of risk factors on hypertension and T2D

Partial LDSC analyses helped understand the potential mechanisms underlying the genetic correlation between T2D and hypertension. This method tests the hypothesis that a genetic correlation remains after accounting for the genetic component captured by genetic data from a risk factor phenotype.

Our partial LDSC analysis found that the genetics of 8 of 10 common risk factors significantly attenuated the genetic correlation between hypertension and T2D, with BMI, waist-hip ratio (WHR), and triglycerides having the biggest effects (Figure 3A). When combining all 10 risk factors in a multivariate analysis, BMI, waist-hip ratio and triglycerides independently attenuated the genetic correlation between hypertension and T2D (Figure 3B): adjusting for additional risk factors did not further attenuate the genetic correlation (Supplementary Table 3). Significant genetic correlation remained after removing the genetic effects of these three risk factors (r_g_ = 0.31, SE = 0.02, p = 2.03*10^-36^), indicating remaining shared causal mechanisms not acting through the genetic components of BMI, WHR, or triglycerides captured by existing GWAS data.

### Shared causal mechanisms before removing the causal effects of common risk factors

To identify specific genetic variants and potential mechanisms behind the shared component to T2D and hypertension, we performed colocalisation analyses. This method allows us to assess the probability that given genomic regions contain a genetic variant that is causal for two traits. We first did this using the original hypertension and T2D summary statistics, prior to adjusting for common shared risk factors.

There were 4,337 SNPs in 98 genomic regions significantly associated with both hypertension and T2D (p < 5*10^-8^). Thirty-three of these regions had evidence of colocalising variants (PPH3+PPH4 ≥ 0.8 and PPH4/PPH3 > 5), with a total of 400 variants in the 95% credible sets, ranging from one to 109 variants per locus (Supplementary Table 4). For 15 of the 33 colocalising regions, a single genetic variant had >50% probability (PPH4) of being the shared causal variant for both hypertension and T2D. The remaining 18 regions contained multiple variants in the 95% credible sets.

All 15 colocalising variants (or highly correlated variants: R^2^>0.9) with PPH4 >50% have previously been associated with T2D [49], seven have previously been associated with hypertension or blood pressure, with nine associated with one or more of: BMI (n=7), WHR (n=4), or cholesterol (n=5) in the GWAS Catalog (Supplementary Table 5). The hypertension risk-increasing allele was also the T2D risk-increasing allele for all variants except rs12921916 (PPH4=1), in the *PDILT* gene, where the T-allele increased risk of hypertension, but decreased risk of T2D. This allele is associated with decreased estimated glomerular filtration rate and risk of chronic kidney disease, and also increased blood urea nitrogen levels, but decreased risk of kidney stone disease (Figure 4).

**Figure 4:**
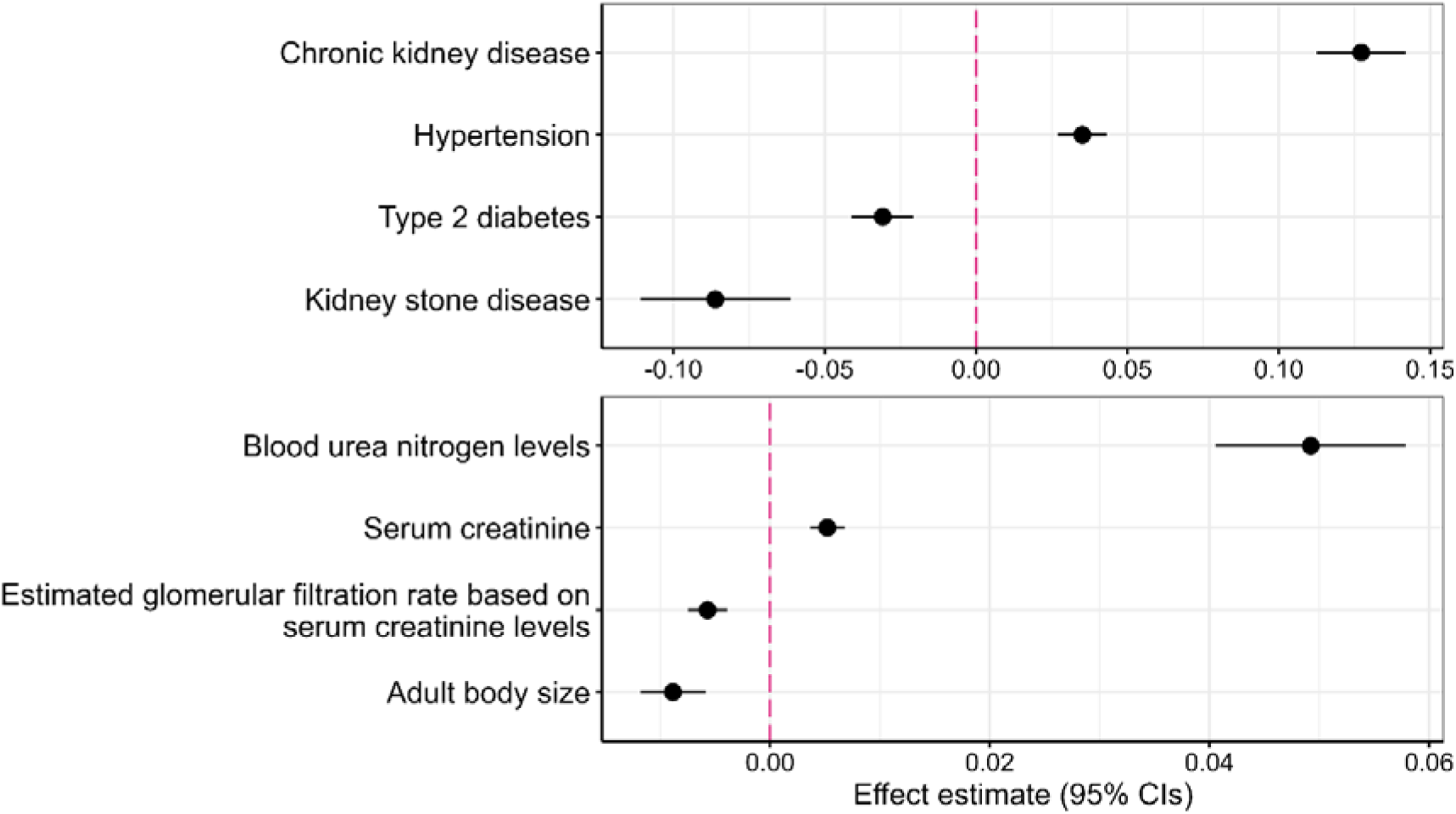
Published associations between rs12921916 in *PDILT* and traits in the GWAS Catalog. Association of rs12921916-T with hypertension and type 2 diabetes in GEMINI hypertension and T2D GWAS summary statistics, and associations with other traits published in the GWAS Catalog [40]. The GWAS Catalog was also searched for proxy variants (R^2^>0.9); trait-associations with these variants are also included in the plot. Date of search was 3^rd^ June 2025. Each variant is coded to align with the hypertension risk-increasing allele. Note that the scales for the effect estimates differ, so a direct comparison between all traits is not possible. For diseases (top), the estimate is the log-odds ratio. For quantitative traits (bottom), it is (usually) the standardised beta. See Supplementary Table 5 for details.

### Shared causal mechanisms after removing the causal effects of common risk factors

We then wanted to investigate whether any shared causal mechanisms remained between hypertension and T2D after removing the causal effects of the common risk factors, BMI, WHR, and triglycerides. After using our genetic approach to remove the causal effects of BMI, WHR, and triglycerides from the GWAS summary statistics for both hypertension and T2D, 1,517 variants were significantly associated with both conditions (p < 5*10^-8^). These variants covered 35 genomic regions. Twelve of these regions had evidence of colocalisation (PPH3+PPH4 ≥ 0.8 and PPH4/PPH3 > 5), with a total of 35 distinct variants in the 95% credible sets, ranging from one to six variants in the credible set at each locus (Supplementary Table 6).

Of the 35 variants in the 95% credible sets, 17 were in the credible sets prior to risk factor adjustment. Of the 12 regions with evidence of colocalisation after risk factor adjustment, eight were amongst the 33 regions identified prior to risk factor adjustment; for six of these regions, the lead variant was the same before and after adjustment. Four regions had evidence of colocalisation after removing the causal effects of BMI, WHR, and triglycerides, but not before: two regions on chromosome 11, one on chromosome 16, and one on chromosome 19.

Of the 12 regions with evidence of colocalisation, we identified a specific variant with >50% probability (PPH4) of being the shared causal variant for eight regions (Table 1). Four regions on chromosome 8 spanning ∼2.8mb had moderate LD (ranging from R^2^ 0.33-0.51) between the lead variants (the variant with the highest probability of being the colocalising variant). To investigate whether these four regions represented independent signals, we used GCTA-COJO[39] to perform a conditional analysis on both the hypertension and T2D risk-factor-adjusted summary statistics. We chose the variant most significantly associated with hypertension (rs10090444) out of the four lead variants. After conditioning on this variant, no other variants of genome-wide significance remained in the region. This variant lies upstream of *PINX1*.

**Table 1:** Independent genetic variants with high probability of being causal for both hypertension and type 2 diabetes in colocalisation analysis after removing the causal effect of obesity and triglycerides.

| Variant | Chromosome and position (GRCh37) | Effect/ Other allele | Hypertension Beta (SE) | T2D Beta (SE) | Nearest gene (protein-coding) | GWAS Catalog associations (this SNP and proxy SNPs, $R^2 > 0.9$) | pQTL? (UKB Olink proteomics data) |
| --- | --- | --- | --- | --- | --- | --- | --- |
| rs10090444 | 8:10745469 | C/G | 0.0431 (0.0039) | 0.0374 (0.0051) | *PINX1* | Systolic blood pressure, endometriosis, BMI | *Trans*-pQTL for 53 proteins |
| rs5215 | 11:17408630 | C/T | 0.0269 (0.0039) | 0.0662 (0.0050) | *KCNJ11* * | T2D, HbA1c, glucose levels, BMI, blood pressure, pulse pressure, hematocrit, blood urea nitrogen | *Cis*-pQTL for one protein, *Trans*-pQTL for 7 proteins |
| rs11216316 | 11:117081500 | C/A | 0.0508 (0.0063) | 0.0569 (0.0079) | *PCSK7* * | Triglyceride levels, soluble transferrin receptor concentration, iron status biomarkers, cholesterol levels | *Cis*-pQTL for one protein, *Trans*-pQTL for 26 proteins |
| rs56408111 | 19:19793545 | C/T | 0.0585 (0.0076) | 0.0948 (0.0109) | *ZNF101* * | Cholesterol levels, BMI, blood pressure, liver enzymes, bilirubin, calcium, vitamin D | *Trans*-pQTL for 55 proteins |

`coloc` analysis resulted in 8 variants with high probability (SNP PPH4>0.9) of being causal for both conditions after `bGWAS` removal of the causal effect of BMI, WHR, and triglycerides. Following GCTA-COJO conditional analysis, we concluded 4 of these were independent signals. See Supplementary Tables 6 and 7.
Nearest gene: \* indicates the variant is within the gene, otherwise the 'nearest' gene is judged by distance to transcription start site.

Similarly for two colocalising regions on chromosome 11, the lead variants were nearby (∼300kb) and in moderate LD (R^2^=0.77). We used GCTA-COJO to condition on the most significant of the two variants (rs11216316). After conditioning on this variant, the second signal was no longer genome-wide significant, indicating it was not independent.

This left us with four independent colocalising regions, where a specific variant was identified as being causal for both hypertension and T2D. For all four variants, the risk-increasing allele for hypertension was the same as for T2D. Three of the four variants have been associated with at least one other trait (p<5*10^-8^) in the GWAS Catalog; one variant had no known associations other than the hypertension and T2D association we identified, but proxy variants (R^2^ > 0.9) had been previously reported as associated with a range of traits, including SBP, BMI and endometriosis (Supplementary Table 7; Figure 5; Supplementary Figures 4, 5, 6).

**Figure 5:**
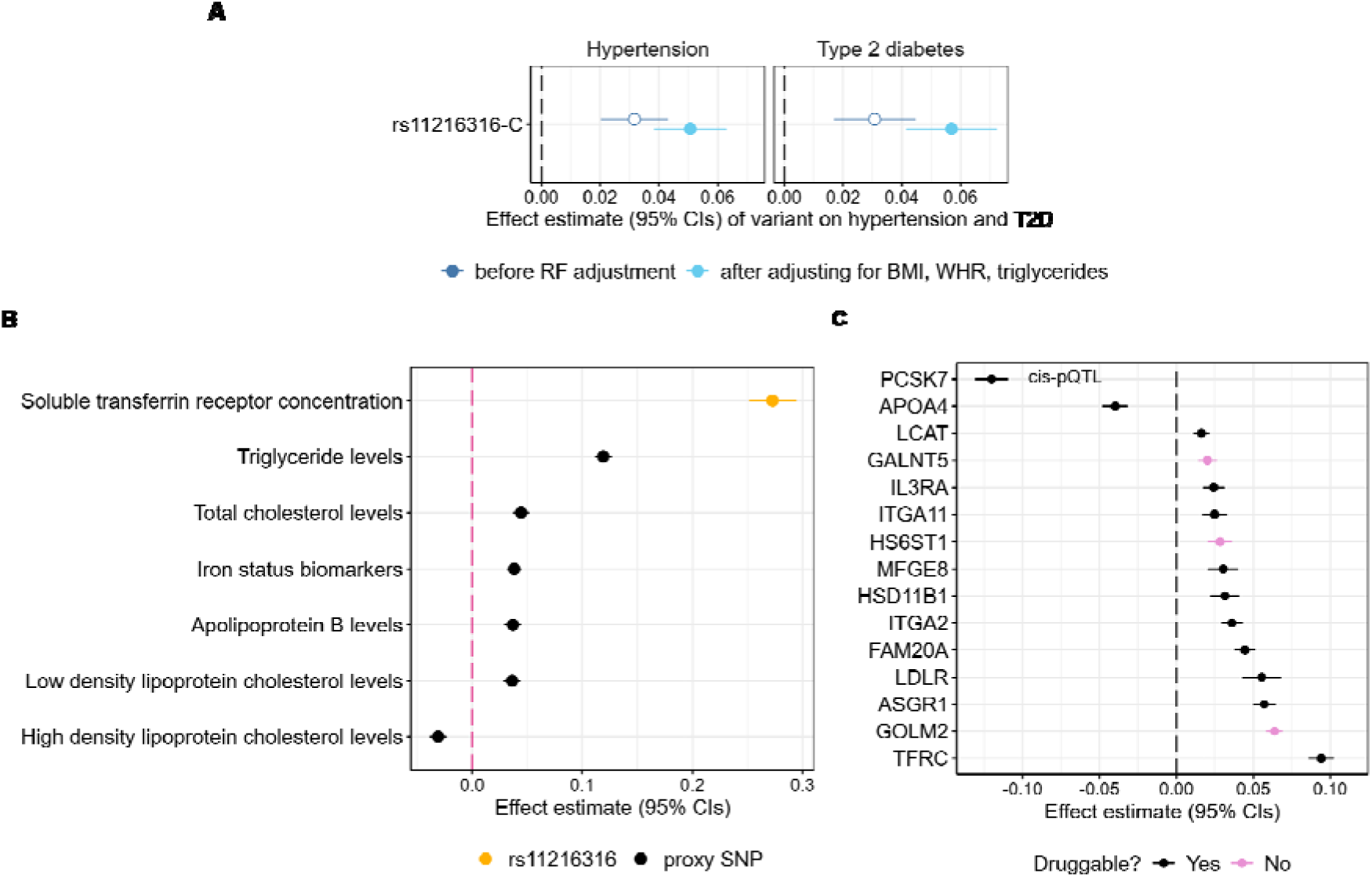
Genetic variant rs11216316 in the *PCSK7* locus associations with known traits and protein levels. A) rs11216316 associations with hypertension and type 2 diabetes in GEMINI data before and after removing the causal effect of BMI, WHR and triglycerides. B) rs11216316 associations with published results from GWAS Catalog (or a proxy variant in LD with R^2^>0.9) [40], see Supplementary Table 7 for full results. C) top 15 most significantly associated proteins with rs11216316 in UK Biobank proteomics data [45], see Supplementary Table 8 for full results.

### Proteomic analysis

To assess the potential of our results in identifying targets for intervention or disease prevention, we performed proteomic analysis to see whether the four variants we had identified in our colocalisation analysis were associated with levels of circulating proteins, and whether those proteins could be targeted by drugs or interventions.

The four variants identified by the colocalisation analysis after removing the causal effects of the three risk factors were associated (p<5*10^-5^) with a total of 143 circulating proteins (some proteins were associated with more than one of the four variants; there were 136 unique proteins) in our proteomics analysis. Of the 136 unique proteins, 101 are classified as druggable in the druggable genome; the drug-gene interaction database showed 47 of those are targeted by at least one approved drug.

Two of the four genetic variants with significant colocalisation between hypertension and T2D after removing the causal effects of BMI, WHR and triglycerides were identified as a *cis*-pQTL in our analysis: rs11216316-C was associated with lower serum PCSK7 levels (Figure 5; Supplementary Table 8) and rs5215-C was associated with lower serum NCR3LG1.

### Observational relationship between hypertension and T2D

To provide some clinical context to our genetic analyses, we conducted observational analyses using real-world health data. N= 3,594,211 patients with hypertension, T2D or both were identified in the CPRD-Aurum database; 65.7% (755,706/1,150,916) of people with T2D also had hypertension, compared to 23.6% (755,706/3,199,001) of people with hypertension having T2D (Figure 6A). The median age at diagnosis was 57.9 (IQR 49.0–66.9) for hypertension and 62.1 (IQR 52.7–71.4) years for T2D. Among patients with both diseases, hypertension was more likely to be diagnosed first (65.7%, 95% CI: 65.6–65.8%). The median interval between diagnoses was 5.8 (IQR 1.9–11.6) years, with a longer disease interval in hypertension-first cases (median 7.5 [IQR 3.1–13.4] years) compared to T2D-first cases (3.9 [IQR 1.2–8.5] years, Figure 6B). 3.7% of patients received both diagnoses on the same day, 7.1% within one month and 17.9% within 12 months. Comorbidity patterns show shared clinical burden: patients with both hypertension and T2D exhibited higher rates of chronic kidney disease (29.9% vs 21.4% overall, p<0.001), obesity (21.9% vs 12.6% overall, p<0.001) and more LTCs at the time of diagnosis (median 6 [IQR 4-9] vs median 5 [IQR 3-8] overall, p<0.001) (Supplementary Table 9).

**Figure 6:**
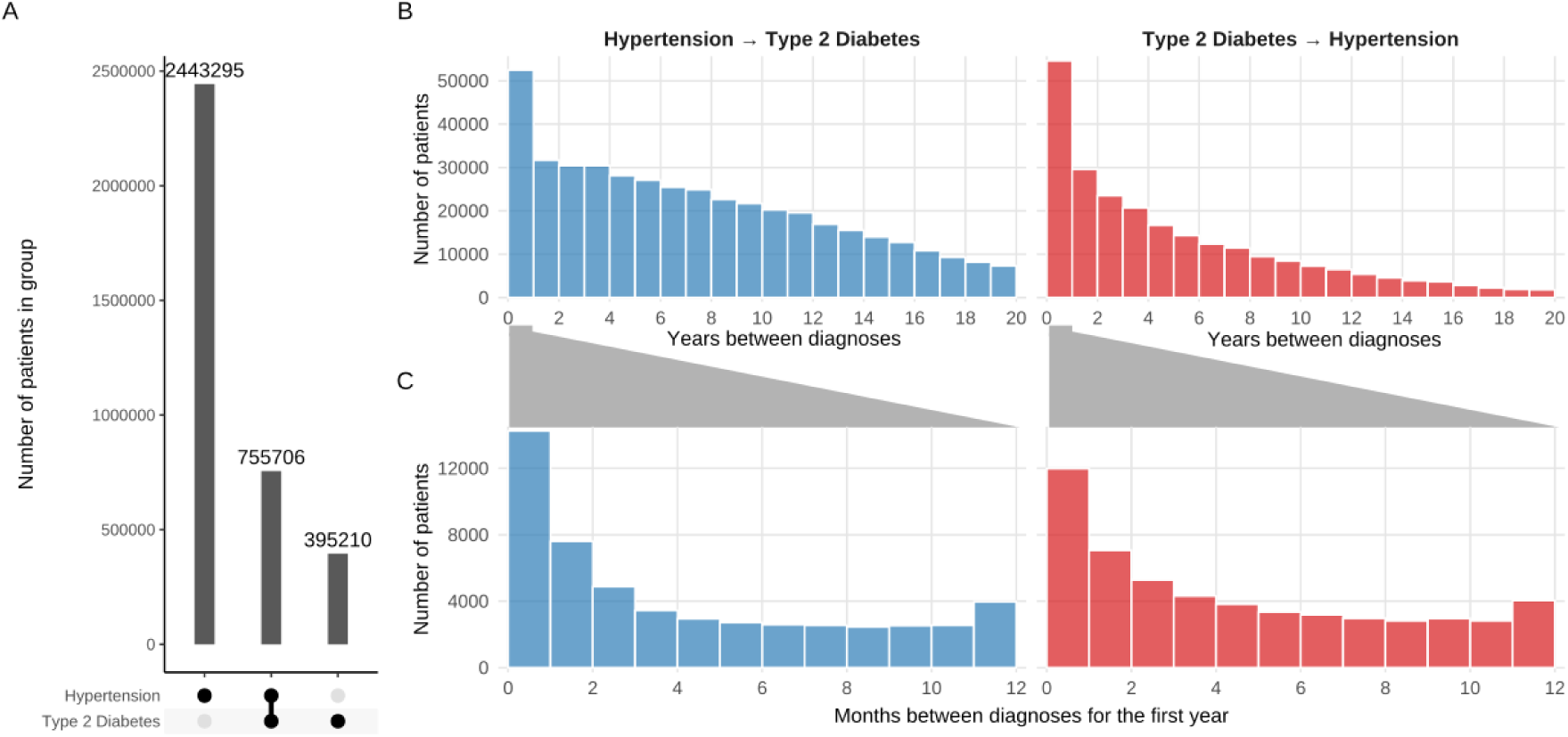
Relationship between hypertension and T2D diagnosis in CPRD. A) Upset plot showing the number of patients diagnosed with either hypertension or T2D, and the number in each group; B) Bar chart showing the time intervals between diagnoses from hypertension to T2D (blue), and T2D to hypertension (red); C) same as B but by month for the first 12 months.

## Discussion

We used large-scale human genetics data sources to generate new insights into the co-occurrence of hypertension and T2D. Importantly, our genetic approach identified shared risk factors that were independent of the established, adiposity-related, risk factors (BMI, fat distribution [WHR], and circulating triglycerides).

Using colocalisation methods, we identified four independent genetic loci, each with specific genetic variants, causal for both hypertension and T2D beyond the traditional modifiable risk factors, BMI, WHR, and triglycerides. Proteins associated with these loci present opportunities for novel drug treatments.

Our key finding highlights a potential opportunity for a novel therapeutic target. We identified rs11216316 in *PCSK7*, with high probability of being causal for both hypertension and T2D. The variant is a *cis*-pQTL for PCSK7 (<22kb from the gene transcription start site), highlighting a possible link between PCSK7 protein levels and increased risk of both hypertension and T2D. PCSK7 is a protein convertase with multiple functions; iron metabolism being a key one. It affects shedding of transferrin receptor protein 1 (TfR1) from the cell surface, influencing serum levels of soluble transferrin receptor (sTfR) [50]. rs11216316 has been identified in GWAS for sTfR [51,52], and is a *trans*-pQTL for TfR1. Iron dysregulation can impact pathways that lead to hypertension or T2D [53,54]; therapeutics that regulate iron homeostasis may help prevention of both hypertension and T2D. More research is needed into the physiological consequences of TfR1 shedding by PCSK7, to confirm whether it indeed impacts iron metabolism – mouse models have been uninformative as while human TfR1 is shed by PCSK7, mouse TfR1 is not, due to species’ differences [50]. The *PCSK7* variant, and a variant in strong LD, have also been associated with circulating lipid levels and metabolic dysfunction-associated steatotic liver disease (MASLD) [55,56], highlighting another potential, and intervenable, path to the development of hypertension and T2D. PCSK7 is not currently the target of any approved drugs; however, it is listed in the druggable genome [46], and its potential as a safe therapeutic target has been proposed recently, particularly for the prevention of MASLD [57,58]. Given that PCSK7 is upstream of several disease-relevant pathways, it is a very attractive target for prevention of hypertension and T2D, along with wider multimorbidity.

A recently published study investigated the shared genetic architecture of T2D and blood pressure (BP), clustering genetic variants into different groups of pathogenetic processes [14]. Methodologically, the two studies are quite distinct: Pascat *et al*. curated a set of variants that were reported genome-wide significant in previous genetic studies for T2D (published between 2012-2018) or for either one of three blood pressure traits (systolic blood pressure, diastolic blood pressure, or pulse pressure; between 2009-2019) – only nine variants were genome-wide significant for both T2D and a blood pressure trait. By contrast, we meta-analysed GWAS data (from 2022 onwards) for hypertension and T2D and used colocalisation to identify variants that were causal for both conditions. Pascat *et al*. clustered variants into distinct groups: metabolic syndrome, higher adiposity, vascular dysfunction, reduced beta-cell function, and inverse T2D-BP risk, highlighting different pathogenetic processes underlying the relationship between T2D and BP. We also explored shared mechanisms between hypertension and T2D, but we instead highlight causal mechanisms that remain after adjusting for the genetic effects of adiposity measures captured by GWAS and explore their potential as therapeutic targets. None of our four variants that colocalised after accounting for adiposity were included in the variants studied by Pascat *et al*., though variants in LD (R^2^ ranging from 0.22-0.60) with three of them (rs10090444, rs5215, rs56408111) were included. This may be because two of our variants (rs56408111, rs11216316) only reached genome-wide significance for both hypertension and T2D after adjusting for the genetic effects of adiposity.

In our analysis, a variant in the 3′ UTR region of gene *ZNF101*, rs56408111, was identified in colocalisation analysis. This variant has been linked to blood pressure and T2D [18,49,59,60], along with MASLD [61]. Genetic variants highly correlated with rs5640811 (corresponding to the hypertension and T2D risk-increasing allele) have been associated with decreased levels of triglycerides, cholesterol, and BMI, but increased levels of liver markers (see Supplementary Table 7). The opposing directions of effect on hypertension/T2D and BMI/WHR/triglycerides may explain why the locus only appeared as causal for hypertension and T2D after removing the genetic effects of BMI, WHR, and triglycerides, and the variant’s relationship with liver markers suggests a link between this variant’s effect on hypertension and T2D and liver function. A moderately correlated variant (rs58542926, R^2^=0.47) in gene *TM6SF2* increases liver fat and plays a key role in the development of MASLD, while having cardioprotective effects [62–64]; the signal we found in *ZNF101* may not be independent to this one or may be acting in a similar way. More research is needed to establish whether these two loci are independent, and thus whether the *ZNF101* locus has potential as a target for intervention for hypertension and T2D, or whether the focus should be on the *TM6SF2* locus.

Colocalisation identified rs5215, a missense variant in gene *KCNJ11,* that increases glucose levels and risk of T2D. The variant rs5215 results in an amino acid substitution (I337V) in protein Kir6.2 – a major subunit of the ATP-sensitive potassium channel in pancreatic beta-cells, which plays a key role in regulating insulin secretion and therefore T2D [65,66]. rs5215 has been linked to hypertension, though the mechanistic pathway is less clear [67]. It is possible that rs5215 influences ATP-sensitive potassium channel activity in other cells of the body, which could impact the cardiovascular system, leading to hypertension [68,69]. There are existing, approved drugs used in the management of both hypertension and T2D that target the ATP-sensitive potassium channels, such as sulfonylureas, a class of drugs used in the management of T2D. Glimepiride (a common sulfonylurea) blocks ATP-sensitive potassium channels, stimulating the secretion of insulin from pancreatic beta-cells, reducing blood glucose levels [70]. It is not reported to have any effects on blood pressure. Diazoxide is primarily used to manage hypoglycaemia in patients with medical conditions causing them to make too much insulin. It activates ATP-sensitive potassium channels in the pancreatic beta-cells, inhibiting the secretion of insulin. Diazoxide also acts as a vasodilator, which results in lower blood pressure; an intravenous form of the drug is used in hypertensive emergencies [71]. However, it can increase the risk of hypotension when used in combination with various other drugs [72]. Further research is needed into the exact role of pancreatic potassium channel proteins (such as Kir6.2) on hypertension and diabetes, along with potential for safe interventions or treatments for both diseases.

The final colocalising variant identified was rs10090444, which is intergenic on chromosome 8, 46kbp upstream of gene *PINX1*. The variant was previously identified for traits including SBP and endometriosis, but the causal gene is unknown. Further work is needed to elucidate the shared target.

Prior to risk factor adjustment, variants in the *UMOD-PDILT* locus had opposite directions of effect for hypertension and T2D. The hypertension risk-increasing allele increases risk of chronic kidney disease (CKD), whilst decreasing risk of calcium oxalate lithiasis (kidney stones) [73,74]. Uromodulin (encoded by the *UMOD* gene) - expressed in the kidneys and excreted in urine – is involved in salt reabsorption and blood pressure control and has a protective effect on urinary tract infections and kidney stone disease [75,76]. This is consistent with central adiposity (that increases T2D risk) increasing the risk of kidney stones [77].

Analysis of primary care data from 755,706 individuals with both hypertension and T2D found that hypertension is commonly diagnosed first (66% of patients), median 7.5 years before T2D; suggesting a large intervention window for prevention of subsequent diabetes. Using large scale genetics resources, we use MR to show evidence supporting a bidirectional causal relationship between hypertension and T2D. A previous MR study in UK Biobank concluded that genetically predisposed T2D was causal for hypertension but found no evidence for hypertension being causal for T2D [12]. Their reported causal effect of T2D on hypertension (OR=1.07; 95% CIs:1.04-1.10; p=3.4×10^-7^) was smaller than we found using larger, meta-analysed cohort data on the same scale (OR=1.23; 95% CIs:1.21-1.25; p=2.26*10^-190^). We also found a statistically significant causal effect of hypertension on liability to T2D (OR=1.32; 95%CI:1.30-1.35; p=2.23*10^-145^). These differences could be partly due to the healthy participant bias in UK Biobank, and greater statistical power in our meta-analysed data. Whilst the validity of results from MR studies relies on the instrumental variable assumptions being satisfied, our results remained consistent in sensitivity analyses, which included outlier removal to account for variants significantly contributing to heterogeneity, and use of more pleiotropy-robust methods. We acknowledge that MR analyses using binary exposures should be interpreted more as a test of the causal null hypothesis, rather than an attempt to calculate an exact causal estimate [24,25].

In the UK, effective chronic disease management, including for hypertension and T2D, and adherence to clinical guidelines is encouraged through financial renumeration for primary care practices (see NHS Quality and Outcome Framework for details https://qof.digital.nhs.uk). Our results suggest the following clinical applications: first, the systematic application of T2D screening tests in hypertensive patients, and vice versa, especially in the early years following diagnosis. In addition, clinical guidelines should be developed that take metabolic and genetic criteria into account. Secondly, genetic correlation and bidirectional causality suggest that treating one disease may improve the other. Thirdly, clinical decision support systems and clinical practice guidelines may need to be updated based on the diagnosis of the first disease. Fourthly, we have identified some important pharmacological intervention targets which need development to reduce disease burden.

Our study has several strengths: we use large UK population-representative patient data to model the patterns of diagnosis, and we leverage large-scale genetic data to identify overlapping causal mechanisms, after subtracting the effects of BMI, WHR and triglycerides. The study has several limitations: first, genetic analyses are restricted to participants of European-like genetic ancestry due to data availability. Second, both hypertension and T2D can go undiagnosed, especially in the early stages, and our analysis is of clinically diagnosed hypertension and T2D [78,79]. Additionally, a diagnosis of one of the conditions may prompt increased interaction and monitoring from healthcare providers, highlighting the presence of the other condition. This may explain why such a high percentage (17.9%) of people with both hypertension and T2D receive both diagnoses within a year.

## Conclusions

Using genetic methods, we identified shared genetic mechanisms between hypertension and T2D independent of established, adiposity-related risk factors (BMI, WHR, and circulating levels of triglycerides). We highlight potential candidate targets for intervention, especially PCSK7. Our findings emphasise the advantage of using a genetic approach to uncover novel opportunities for prevention and treatment optimisation, including drug repurposing or development. As the population ages and more people experience multiple long-term health conditions, it is important that we improve our understanding of disease co-occurrence. Genetic approaches such as the one used here for hypertension and T2D, are key to accomplishing this.

## Supporting information

STROBE MR checklist

## Abbreviations

eQTL: expression quantitative trait loci
GWAS: genome-wide association study
IV: instrumental variable
IVW: inverse-variance weighted
LD: linkage disequilibrium
LDSC: linkage disequilibrium score regression
MASLD: metabolic dysfunction-associated steatotic liver disease
MLTC: multiple long-term conditions
MR: Mendelian randomisation
pQTL: protein quantitative trait loci
SNP: single nucleotide polymorphism
T2D: type 2 diabetes
WHR: waist-hip ratio

## Declarations

### Ethics approval and consent to participate

Data from the Clinical Practice Research Datalink (CPRD) was obtained under license from the UK Medicines and Healthcare products Regulatory Agency. The data is provided by patients and collected by the NHS as part of their care and support. Approval for CPRD data access was granted by the CPRD Independent Scientific Advisory Committee, protocol number 23_003109. The Northwest Multi-Centre Research Ethics Committee approved the collection and use of UK Biobank data for health-related research (Research Ethics Committee reference 11/NW/0382). Access to UK Biobank data was granted under UK Biobank approved application 14631.

### Consent for publication

Not applicable.

### Availability of data and materials

Most data generated during this study are included in this published article and its supplementary information files. We cannot make individual-level data available. Researchers can apply to UK Biobank (https://www.ukbiobank.ac.uk/enable-your-research/) and CPRD (https://www.cprd.com/research-applications). We have made our diagnostic code lists, scripts, and results available on our GitHub (https://github.com/GEMINI-multimorbidity/).

### Authors’ Contributions

Funding acquisition: JD, CV, SK, CF, SEL, MM, LF, KB, FD, JB, TMF, JAHM, and LCP. Conceptualisation: TMF, JAHM, and LCP. Project administration and supervision: JD, TMF, JAHM, and LCP. Data curation: BV, RMW, QG, OM, LCP. Methodology: BV, RMW, QG, OM, FD, JB, TMF, JAHM, LCP. Resources: LCP. Formal analysis, investigation, and software: BV, RMW, and QG. Visualisation: BV, QG. Writing - original draft: BV, RMW, QG, TMF, JAHM, LCP. Writing - review and editing: BV, RMW, QG, OM, JD, CV, SK, DT, CF, SEL, MM, LF, KB, FD, JB, TMF, JAHM, LCP. All authors read and approved the final manuscript.

## Acknowledgments

This is a summary of independent research conducted at the National Institute for Health and Care Research (NIHR) Exeter Biomedical Research Centre (BRC) and by the NIHR Leicester BRC. The work was funded by the UK Medical Research Council (MRC). The interpretation and conclusions contained in this study are those of the authors and not necessarily those of the MRC, NIHR or the Department of Health and Social Care, nor of any other funders. This research has been conducted using the UK Biobank Resource, under application 14631. The authors would like to acknowledge and thank the participants and investigators of the UK Biobank and the FinnGen study. The authors would like to acknowledge both past and present members of the GEMINI Consortium. We acknowledge the use of the University of Exeter High-Performance Computing (HPC) facility in carrying out this work.

For the purpose of open access, the author has applied a Creative Commons Attribution (CC BY) license to any Author Accepted Manuscript version arising from this submission.

