## Supplementary material for "Genome-wide association studies identify shared mechanisms between hypertension and type 2 diabetes independent of adiposity": STROBE MR checklist

### STROBE-MR checklist of recommended items to address in reports of Mendelian randomization studies[1 2]

| Item No. | Section | Checklist item | Page No. | Relevant text from manuscript |
|---|---|---|---|---|
| 1 | **TITLE and ABSTRACT** | Indicate Mendelian randomization (MR) as the study's design in the title and/or the abstract if that is a main purpose of the study | 2 | Not in title as not the main purpose of the study. Mentioned in abstract: 'We investigated the bidirectional causal relationship using Mendelian randomisation'. |
| | **INTRODUCTION** | | | |
| 2 | **Background** | Explain the scientific background and rationale for the reported study. What is the exposure? Is a potential causal relationship between exposure and outcome plausible? Justify why MR is a helpful method to address the study question | 5 | Introduction paragraph 3: 'In this study, we investigate the shared genetic architecture underlying the co-occurrence of hypertension and T2D. We know that the two conditions are genetically correlated [1]: here we assess whether there are bidirectional causal relationships using Mendelian randomisation, a statistical causal inference method that (under specific assumptions) uses genetic variants associated with a trait of interest as instrumental variables to investigate whether the trait has a causal effect on a second trait. We then explore the genetic contribution of common, modifiable risk factors to the genetic relationship, and uncover shared genetic mechanisms acting through pathways distinct to known risk factors. Understanding the mechanistic overlap between this frequently co-occurring condition pair could uncover pathways for prevention and support treatment optimisation, including drug discovery.' |
| 3 | **Objectives** | State specific objectives clearly, including pre-specified causal hypotheses (if any). State that MR is a method that, under specific assumptions, intends to estimate causal effects | 5 | Introduction paragraph 3—same as above. |
| | **METHODS** | | | |
| 4 | **Study design and data sources** | Present key elements of the study design early in the article. Consider including a table listing sources of data for all phases of the study. For each data source contributing to the analysis, describe the following: | | |
| | a) | Setting: Describe the study design and the underlying population, if possible. Describe the setting, locations, and relevant dates, including periods of recruitment, exposure, follow-up, and data collection, when available. | 6 | Methods: Data and sources, paragraph 2: 'To maximise genetic sample sizes when analysing each condition, we meta-analysed genome-wide association study (GWAS) data from three |

| | | | | |
|---|---|---|---|---|
| | | | | sources: two large cohort studies UK Biobank (N=502,000), FinnGen (release 9; N= 377,000) [16] and condition-specific consortium data when available [1]. Analyses only included participants genetically similar to the 1000 Genomes European (EUR) subset' |
| | b) | Participants: Give the eligibility criteria, and the sources and methods of selection of participants. Report the sample size, and whether any power or sample size calculations were carried out prior to the main analysis | 6 | Methods: Data and sources, paragraph 3: 'For hypertension, we meta-analysed GWAS from UK Biobank (N diagnosed=158,746, N controls=291,451) and FinnGen (N diagnosed=111,581, N controls=280,842). No public GWAS consortia meta-analysis was available at the time. For T2D, we meta-analysed a published GWAS consortium meta-analysis (EUR subset N cases=80,154, N controls=853,816) [18] with data from FinnGen (N diagnosed=57,698, N controls=450,121) (the consortium GWAS already included UK Biobank). This gave effective sample sizes for meta-analysis of 725,376 for hypertension and 446,142 for T2D.' |
| | c) | Describe measurement, quality control and selection of genetic variants | 8 | Methods: The genetic relationship between hypertension and type 2 diabetes, paragraph 3: 'To select the genetic variants as instrumental variables for the exposure, we identified the genome-wide significant variants (p < 5*10-8), and estimated independent genetic loci based on a distance of 500kb using the function "get_loci" from the R-package "gwasRtools" (version 0.1.7, https://github.com/lcpilling/gwasRtools). The SNP with the smallest p-value in each locus was labelled as the lead SNP; these were filtered to those present in the outcome GWAS summary statistics and selected as IVs.' |
| | d) | For each exposure, outcome, and other relevant variables, describe methods of assessment and diagnostic criteria for diseases | 6 | Diagnostic code lists used to generate genome-wide association study summary statistics data are described in previous study: 'The first GEMINI study ascertained 72 common, chronic long-term conditions with significant heritability [1]. See GitHub for curated diagnostic code lists (https://github.com/GEMINI-multimorbidity)' |
| | e) | Provide details of ethics committee approval and participant informed consent, if relevant | 14 | Methods: Ethics statement: 'Individual level UK Biobank genetic data was used for investigating genetic variant co-inheritance and Linkage |

| | | | | | |
|---|---|---|---|---|---|
| | | | | | Disequilibrium in colocalisation and conditional analyses. The Northwest Multi-Centre Research Ethics Committee approved the collection and use of UK Biobank data (Research Ethics Committee reference 11/NW/0382). Participants gave informed consent for the use of their data, health records, and biological materials for health-related research purposes. Access to UK Biobank was granted under Application Number 14631.' |
| 5 | **Assumptions** | | Explicitly state the three core IV assumptions for the main analysis (relevance, independence and exclusion restriction) as well assumptions for any additional or sensitivity analysis | 8<br>Supplementary Information | Methods: The genetic relationship between hypertension and type 2 diabetes, paragraph 5: 'The validity of findings from MR analyses relies on three IV assumptions being satisfied (IV1: relevance, IV2: independence, and IV3: exclusion-restriction; illustrated in Supplementary Figure 1), and two of these cannot be fully empirically tested (IV2 and IV3).'<br>Supplementary Figure 1. |
| 6 | **Statistical methods: main analysis** | | Describe statistical methods and statistics used | | |
| | | a) | Describe how quantitative variables were handled in the analyses (i.e., scale, units, model) | - | Only binary traits were used (hypertension and type 2 diabetes disease status). |
| | | b) | Describe how genetic variants were handled in the analyses and, if applicable, how their weights were selected | 9 | Methods: The genetic relationship between hypertension and type 2 diabetes, paragraph 5: 'We used Cochrane's Q-statistic, a measure of heterogeneity, to test for global pleiotropy [30]. Where heterogeneity was detected (Q > number of IVs –1), we identified any outlying variants using Radial MR. Here, each variant's contribution to Cochrane's Q-statistic is quantified, and outliers are detected using a significance threshold of 0.05 [31]. The exposure-outcome association was re-estimated following removal of any outliers. Steiger filtering was used to assess the validity of the IVs by identifying and removing variants that had a stronger association with the outcome than the exposure [32].' |
| | | c) | Describe the MR estimator (e.g. two-stage least squares, Wald ratio) and related statistics. Detail the included covariates and, in case of two-sample MR, whether the same covariate set was used for adjustment in the two samples | 7-8 | Methods: The genetic relationship between hypertension and type 2 diabetes: 'We performed two-sample MR, using the TwoSampleMR R-package (version 0.6.6) [22], focusing on the |

| | | | | |
|---|---|---|---|---|
| | | | | inverse-variance weighted (IVW) estimate for the primary analysis.’ |
| | | d) Explain how missing data were addressed | 8 | Methods: The genetic relationship between hypertension and type 2 diabetes, paragraph 3: ‘The SNP with the smallest p-value in each locus was labelled as the lead SNP; these were filtered to those present in the outcome GWAS summary statistics and selected as IVs.’ |
| | | e) If applicable, indicate how multiple testing was addressed | | Not applicable. |
| 7 | **Assessment of assumptions** | Describe any methods or prior knowledge used to assess the assumptions or justify their validity | 8-9 | Methods: The genetic relationship between hypertension and type 2 diabetes, paragraph 5: ‘We performed extensive sensitivity analyses to investigate robustness of findings to variations in model assumptions, including effects of pleiotropy (balanced and unbalanced). Relaxing the exclusion-restriction assumption (the IVs are only associated with the outcome through the exposure), we looked for consistency of the IVW estimate with the MR-Egger, weighted median and weighted mode estimates; these are slightly more robust to pleiotropy [26–28]. We included the recently proposed MR-GRIP method, which is invariant to allele coding, unlike MR-Egger [29]. We used Cochrane’s Q-statistic, a measure of heterogeneity, to test for global pleiotropy [30]. Where heterogeneity was detected (Q > number of IVs –1), we identified any outlying variants using Radial MR. Here, each variant’s contribution to Cochrane’s Q-statistic is quantified, and outliers are detected using a significance threshold of 0.05 [31]. The exposure-outcome association was re-estimated following removal of any outliers. Steiger filtering was used to assess the validity of the IVs by identifying and removing variants that had a stronger association with the outcome than the exposure [32].’ |
| 8 | **Sensitivity analyses and additional analyses** | Describe any sensitivity analyses or additional analyses performed (e.g. comparison of effect estimates from different approaches, independent replication, bias analytic techniques, validation of instruments, simulations) | 8-9 | Methods: The genetic relationship between hypertension and type 2 diabetes, paragraph 5—as above. |
| 9 | **Software and pre-registration** | | | |

| | | | | | |
|---|---|---|---|---|---|
| | | a) | Name statistical software and package(s), including version and settings used | 7-8 | Methods: The genetic relationship between hypertension and type 2 diabetes, paragraph 2: 'We performed two-sample MR, using the TwoSampleMR R-package (version 0.6.6) [22]'. |
| | | b) | State whether the study protocol and details were pre-registered (as well as when and where) | | Study was not pre-registered. |
| | **RESULTS** | | | | |
| 10 | **Descriptive data** | | | | |
| | | a) | Report the numbers of individuals at each stage of included studies and reasons for exclusion. Consider use of a flow diagram | 6 | Methods: Data and sources, paragraph 3: 'For hypertension, we meta-analysed GWAS from UK Biobank (N diagnosed=158,746, N controls=291,451) and FinnGen (N diagnosed=111,581, N controls=280,842). No public GWAS consortia meta-analysis was available at the time. For T2D, we meta-analysed a published GWAS consortium meta-analysis (EUR subset N cases=80,154, N controls=853,816) [18] with data from FinnGen (N diagnosed=57,698, N controls=450,121) (the consortium GWAS already included UK Biobank). This gave effective sample sizes for meta-analysis of 725,376 for hypertension and 446,142 for T2D.' |
| | | b) | Report summary statistics for phenotypic exposure(s), outcome(s), and other relevant variables (e.g. means, SDs, proportions) | | Reported in our original, referenced publication: doi:10.1016/j.ebiom.2025.105584. |
| | | c) | If the data sources include meta-analyses of previous studies, provide the assessments of heterogeneity across these studies | | Reported in our original, referenced publication: doi:10.1016/j.ebiom.2025.105584. |
| | | d) | For two-sample MR:<br>i. Provide justification of the similarity of the genetic variant-exposure associations between the exposure and outcome samples<br>ii. Provide information on the number of individuals who overlap between the exposure and outcome studies | 8 | i) Methods: The genetic relationship between hypertension and type 2 diabetes, paragraph 3: 'Both the exposure and outcome GWAS were of individuals of European ancestry, minimising the risk of population stratification and differences in LD structure. Both included adults of similar age and sex distributions, reducing the likelihood of context-dependent genetic effects.'<br>ii) Methods: 'There is therefore substantial sample overlap (>50%) as UK Biobank |

| | | | | | |
|---|---|---|---|---|---|
| | | | | | and FinnGen are included in both GWAS.’ |
| 11 | **Main results** | | | | |
| | | a) | Report the associations between genetic variant and exposure, and between genetic variant and outcome, preferably on an interpretable scale | Supplementary Tables | Reported in Supplementary Table 1. |
| | | b) | Report MR estimates of the relationship between exposure and outcome, and the measures of uncertainty from the MR analysis, on an interpretable scale, such as odds ratio or relative risk per SD difference | 15-16 | Results: Estimated causal effect of hypertension on T2D and Estimated causal effect of T2D on hypertension. |
| | | c) | If relevant, consider translating estimates of relative risk into absolute risk for a meaningful time period | | NA |
| | | d) | Consider plots to visualize results (e.g. forest plot, scatterplot of associations between genetic variants and outcome versus between genetic variants and exposure) | 41<br>Supplementary Information | Figure 2 – bidirectional MR results.<br>Supplementary Figures 2A, 2B, 2B – scatter plots, Supplementary Figure 3 – forest plot of sensitivity analysis results. |
| 12 | **Assessment of assumptions** | | | | |
| | | a) | Report the assessment of the validity of the assumptions | Supplementary Information | Supplementary Results: Estimated causal effect of hypertension on T2D – sensitivity analyses, Estimated causal effect of T2D on hypertension – sensitivity analyses. |
| | | b) | Report any additional statistics (e.g., assessments of heterogeneity across genetic variants, such as $I^2$, Q statistic or E-value) | Supplementary Information | Supplementary Results: Estimated causal effect of hypertension on T2D – sensitivity analyses, Estimated causal effect of T2D on hypertension – sensitivity analyses. |
| 13 | **Sensitivity analyses and additional analyses** | | | | |
| | | a) | Report any sensitivity analyses to assess the robustness of the main results to violations of the assumptions | Supplementary Information, Supplementary Tables | Supplementary Results: Estimated causal effect of hypertension on T2D – sensitivity analyses, Estimated causal effect of T2D on hypertension – sensitivity analyses.<br>Supplementary Figures 2A, 2B, 2C – scatter plots, Supplementary Figure 3 – forest plot of sensitivity analysis results.<br>Supplementary Tables – Supplementary Table 1. |

| | | | | | |
|---|---|---|---|---|---|
| | | b) | Report results from other sensitivity analyses or additional analyses | Supplementary Information, Supplementary Tables | Same as above. |
| | | c) | Report any assessment of direction of causal relationship (e.g., bidirectional MR) | 15-16<br>Supplementary Information.<br>Supplementary Tables. | Results: Estimated causal effect of hypertension on T2D and Estimated causal effect of T2D on hypertension.<br>Figure 2.<br>Supplementary Figure 3 – forest plot of sensitivity analysis results.<br>Supplementary Tables – Supplementary Table 1. |
| | | d) | When relevant, report and compare with estimates from non-MR analyses | 20 | Results: Observational relationship between hypertension and T2D. |
| | | e) | Consider additional plots to visualize results (e.g., leave-one-out analyses) | Supplementary Information. | Scatter plots in Supplementary Figures 2A, 2B, 2C. |
| | **DISCUSSION** | | | | |
| 14 | **Key results** | | Summarize key results with reference to study objectives | 24-25 | Discussion, paragraph 9: 'Using large scale genetics resources, we use MR to show evidence supporting a bidirectional causal relationship between hypertension and T2D. A previous MR study in UK Biobank concluded that genetically predisposed T2D was causal for hypertension but found no evidence for hypertension being causal for T2D [12]. Their reported causal effect of T2D on hypertension (OR=1.07; 95% CIs:1.04-1.10; p=3.4×10-7) was smaller than we found using larger, meta-analysed cohort data on the same scale (OR=1.23; 95% CIs:1.21-1.25; p=2.26*10-190). We also found a statistically significant causal effect of hypertension on liability to T2D (OR=1.32; 95%CI:1.30-1.35; p=2.23*10-145).' |
| 15 | **Limitations** | | Discuss limitations of the study, taking into account the validity of the IV assumptions, other sources of potential bias, and imprecision. Discuss both direction and magnitude of any potential bias and any efforts to address them | 25 | Discussion, paragraph 9: 'These differences could be partly due to the healthy participant bias in UK Biobank, and greater statistical power in our meta-analysed data. Results remained consistent in sensitivity analyses, including outlier removal to account for variants significantly contributing to heterogeneity, and use of more pleiotropy-robust methods. We acknowledge that MR analyses using binary exposures should be interpreted |

| | | | | |
|---|---|---|---|---|
| | | | | more as a test of the causal null hypothesis, rather than an attempt to calculate an exact causal estimate [24,25].’<br>Discussion, paragraph 11: ‘The study has several limitations: first, genetic analyses are restricted to participants of European-like genetic ancestry due to data availability. Second, both hypertension and T2D can go undiagnosed, especially in the early stages, and our analysis is of clinically diagnosed hypertension and T2D [77,78]. Additionally, a diagnosis of one of the conditions may prompt increased interaction and monitoring from healthcare providers, highlighting the presence of the other condition. This may explain why such a high percentage (17.9%) of people with both hypertension and T2D receive both diagnoses within a year.’ |
| 16 | **Interpretation** | | | |
| | | a) Meaning: Give a cautious overall interpretation of results in the context of their limitations and in comparison with other studies | 24-25 | Discussion, paragraph 9: ‘Using large scale genetics resources, we use MR to show evidence supporting a bidirectional causal relationship between hypertension and T2D. A previous MR study in UK Biobank concluded that genetically predisposed T2D was causal for hypertension but found no evidence for hypertension being causal for T2D [12]. Their reported causal effect of T2D on hypertension (OR=1.07; 95% CIs:1.04-1.10; p=3.4×10-7) was smaller than we found using larger, meta-analysed cohort data on the same scale (OR=1.23; 95% CIs:1.21-1.25; p=2.26*10-190). We also found a statistically significant causal effect of hypertension on liability to T2D (OR=1.32; 95%CI:1.30-1.35; p=2.23*10-145).’ |
| | | b) Mechanism: Discuss underlying biological mechanisms that could drive a potential causal relationship between the investigated exposure and the outcome, and whether the gene-environment equivalence assumption is reasonable. Use causal language carefully, clarifying that IV estimates may provide causal effects only under certain assumptions | 21-25 | Discussion, paragraphs 3-9. |
| | | c) Clinical relevance: Discuss whether the results have clinical or public policy relevance, and to what extent they inform effect sizes of possible interventions | 25 | Discussion, paragraph 10. |

| | | | | |
|---|---|---|---|---|
| 17 | **Generalizability** | Discuss the generalizability of the study results (a) to other populations, (b) across other exposure periods/timings, and (c) across other levels of exposure | 26 | Discussion, paragraph 11. |
| | **OTHER INFORMATION** | | | |
| 18 | **Funding** | Describe sources of funding and the role of funders in the present study and, if applicable, sources of funding for the databases and original study or studies on which the present study is based | 29 | Funding. |
| 19 | **Data and data sharing** | Provide the data used to perform all analyses or report where and how the data can be accessed, and reference these sources in the article. Provide the statistical code needed to reproduce the results in the article, or report whether the code is publicly accessible and if so, where | 28 | Availability of data and materials. |
| 20 | **Conflicts of Interest** | All authors should declare all potential conflicts of interest | 28 | Competing interests. |

This checklist is copyrighted by the Equator Network under the Creative Commons Attribution 3.0 Unported (CC BY 3.0) license.

1. Skrivankova VW, Richmond RC, Woolf BAR, Yarmolinsky J, Davies NM, Swanson SA, et al. Strengthening the Reporting of Observational Studies in Epidemiology using Mendelian Randomization (STROBE-MR) Statement. JAMA. 2021;under review.

2. Skrivankova VW, Richmond RC, Woolf BAR, Davies NM, Swanson SA, VanderWeele TJ, et al. Strengthening the Reporting of Observational Studies in Epidemiology using Mendelian Randomisation (STROBE-MR): Explanation and Elaboration. BMJ. 2021;375:n2233.
