## Supplementary Information for "Genome-wide association studies identify shared mechanisms between hypertension and type 2 diabetes independent of adiposity"

### Using Genetics to Understand Shared Mechanisms Between Hypertension and Type 2 Diabetes Accounting for Adiposity

*Voller et al. 2025*

### Supplementary Figures

*Supplementary Figure 1: Mendelian randomisation assumptions*

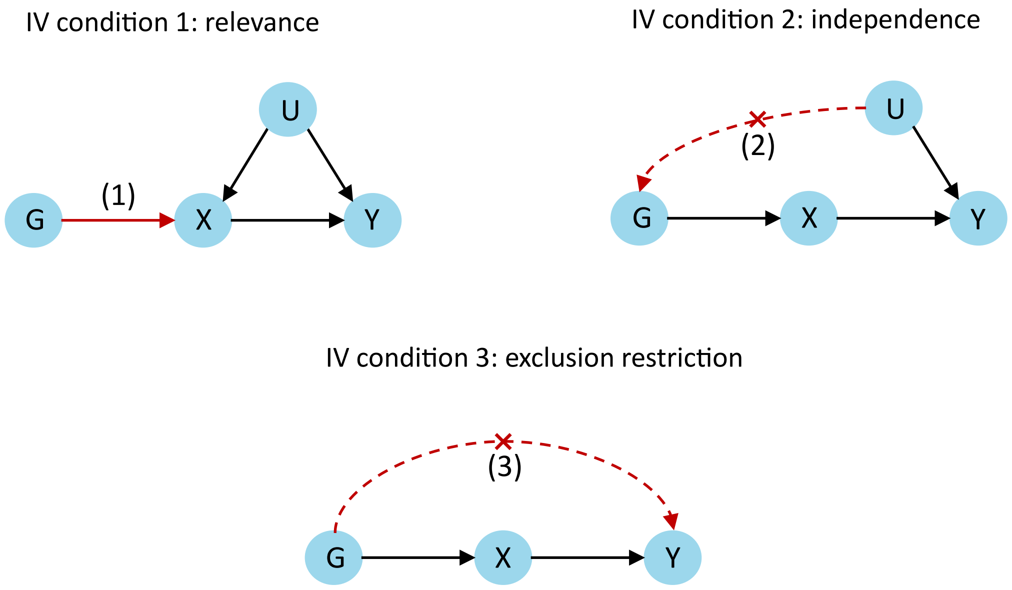

Directed acyclic graphs (DAGs) showing the instrumental variable (IV) conditions; these are required to hold for valid Mendelian randomisation analyses investigating a causal effect of an exposure X on an outcome Y. The conditions are: (1) relevance—the IVs (a set of genetic variants denoted by G) must be associated with the exposure. (2) Independence—there are no causes of G that also influence the outcome Y not via the exposure X (i.e. there are no confounders (U) of the IV-outcome association). (3) Exclusion restriction—the IVs only affect the outcome via the exposure; there is no independent pathway between G and Y (there is no horizontal pleiotropy). Figure adapted from [1].

*Supplementary Figure 2A: MR scatter plots*

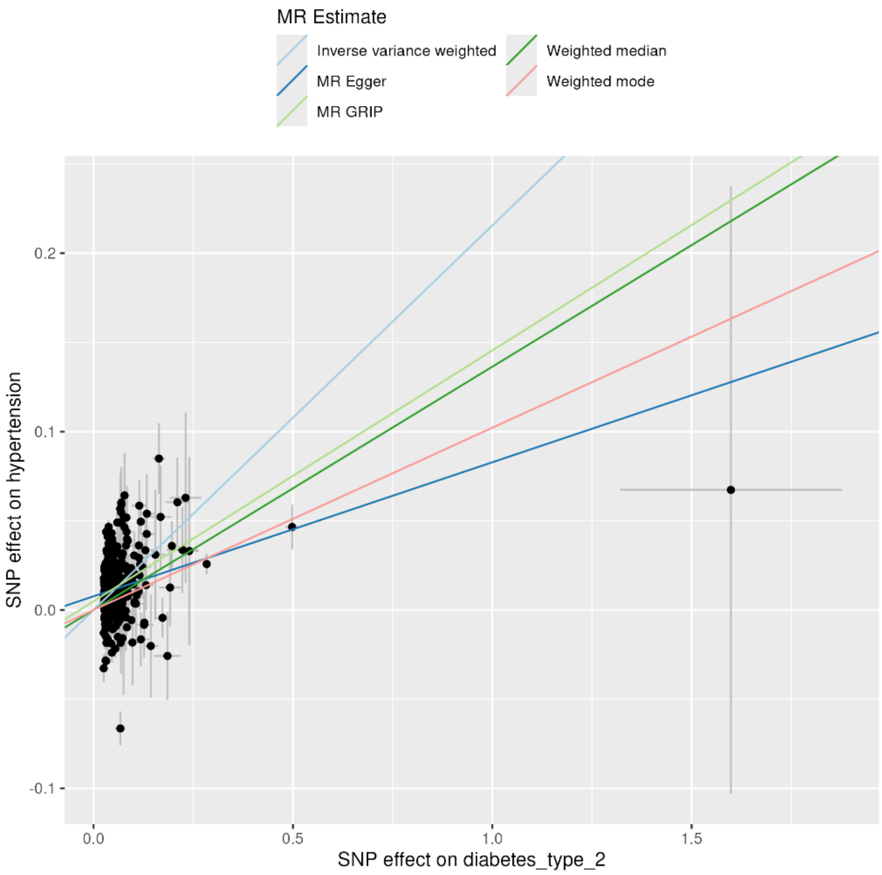

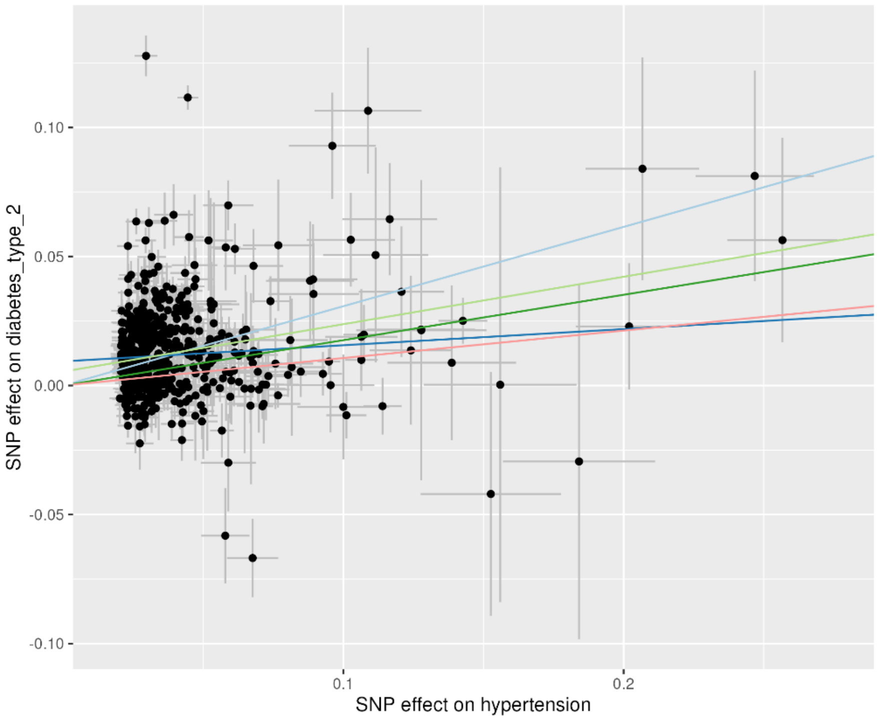

Effect estimates for all SNPs identified as instrumental variables for the MR analyses of the exposure (x-axis) on the outcome (y-axis).

*Supplementary Figure 2B: MR scatter plots after outlier removal*

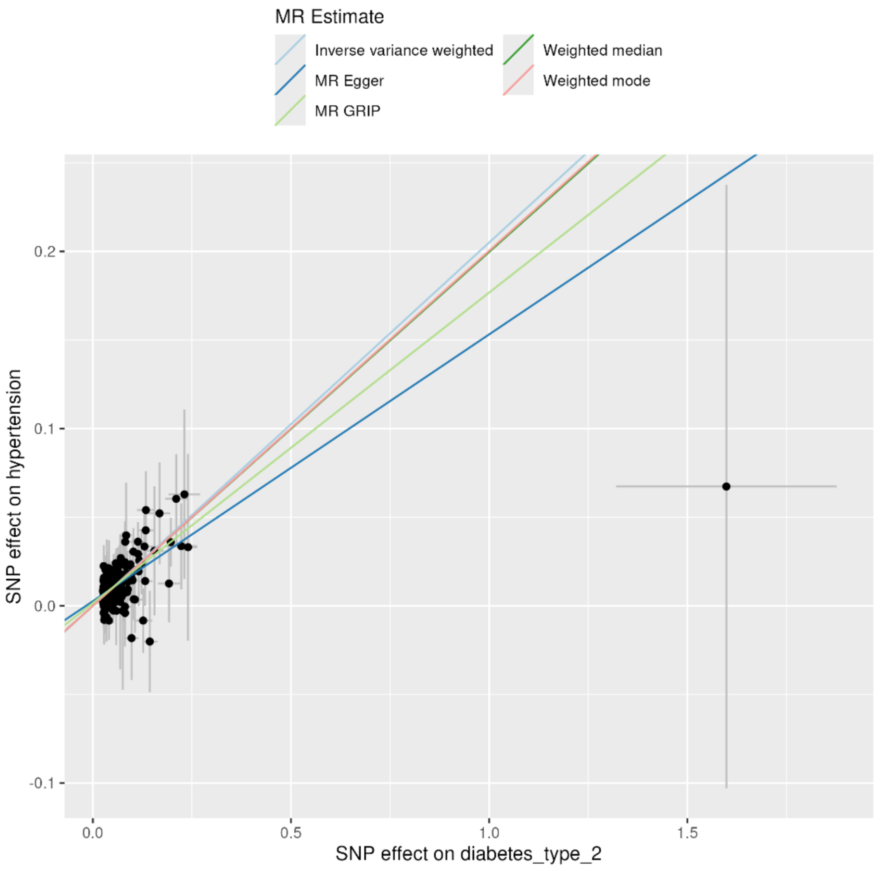

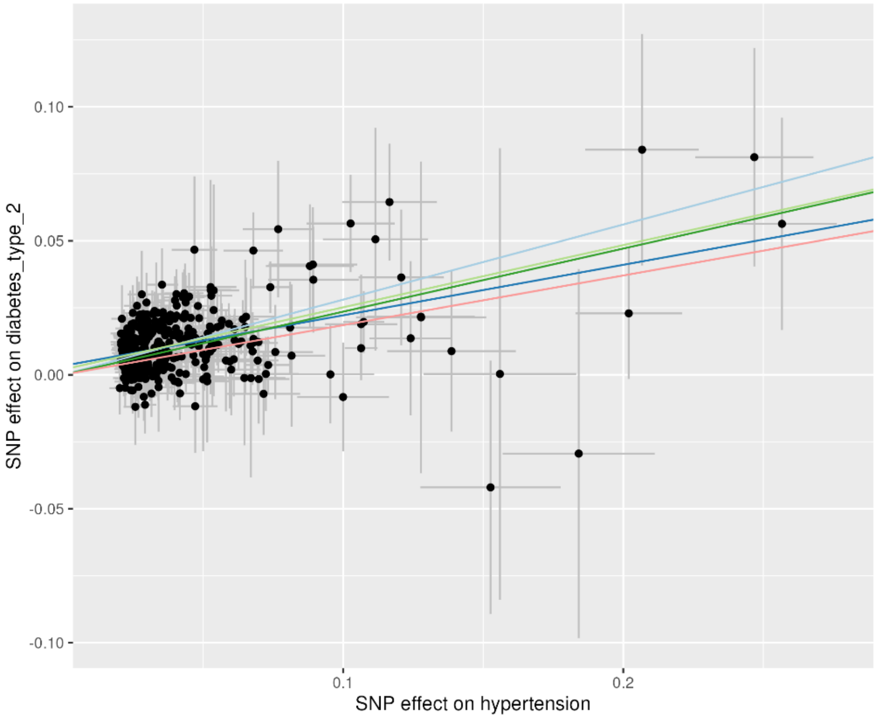

Effect estimates for all SNPs identified as instrumental variables for the MR analyses of the exposure (x-axis) on the outcome (y-axis) following outlier identification (using Radial MR) and removal.

*Supplementary Figure 2C: MR scatter plots after outlier removal and Steiger filtering*

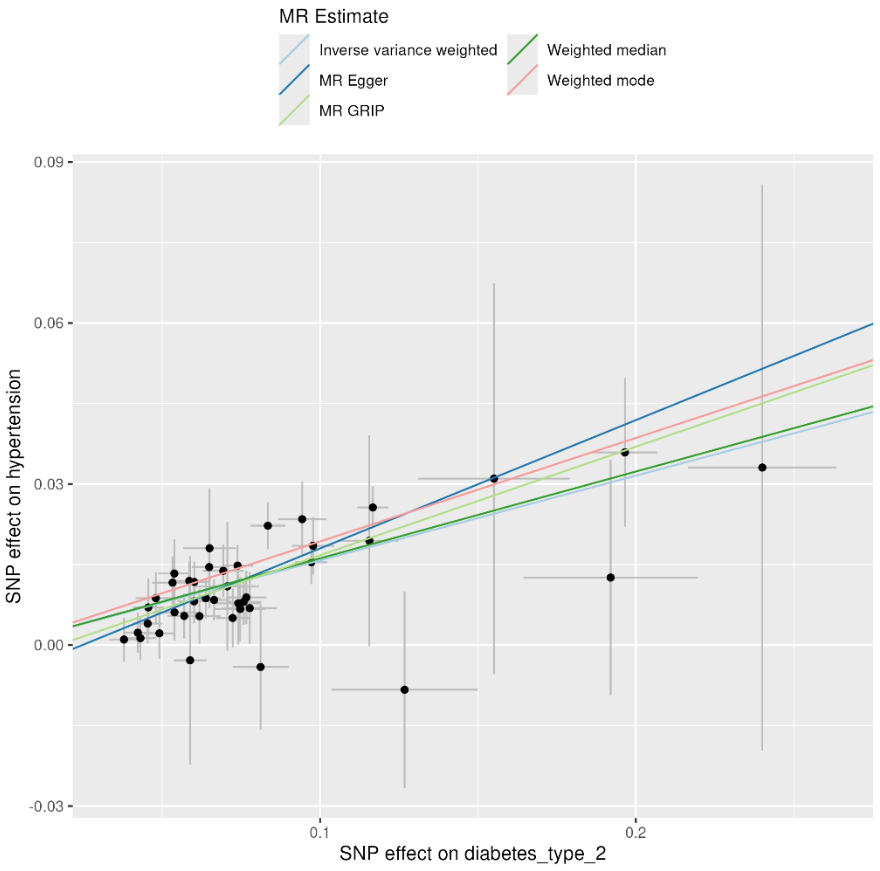

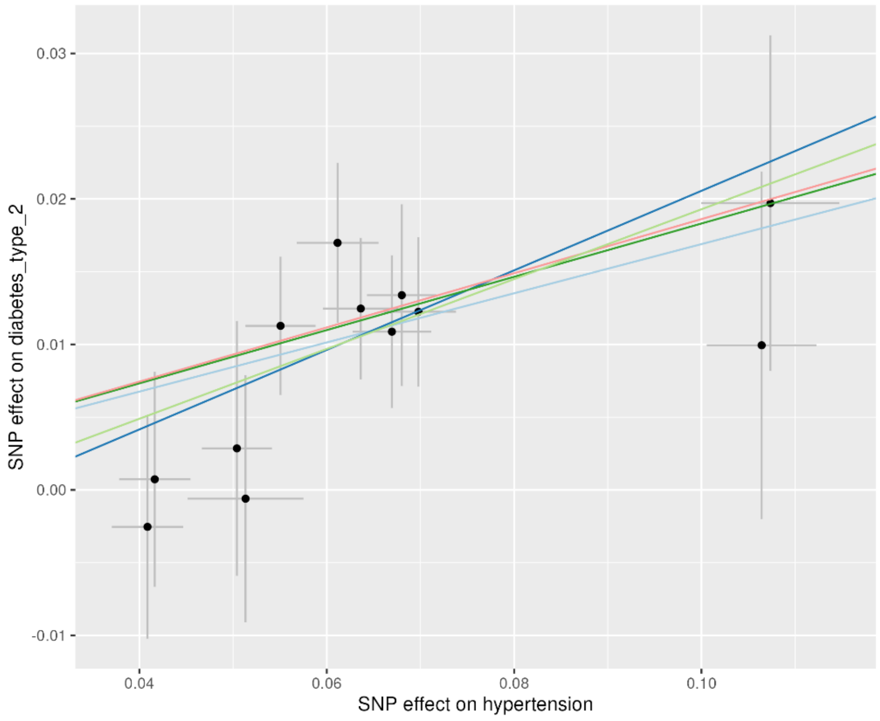

Effect estimates for all SNPs identified as instrumental variables for the MR analyses of the exposure (x-axis) on the outcome (y-axis) following outlier identification (using Radial MR) and removal, and Steiger filtering.

*Supplementary Figure 3: MR sensitivity analysis*

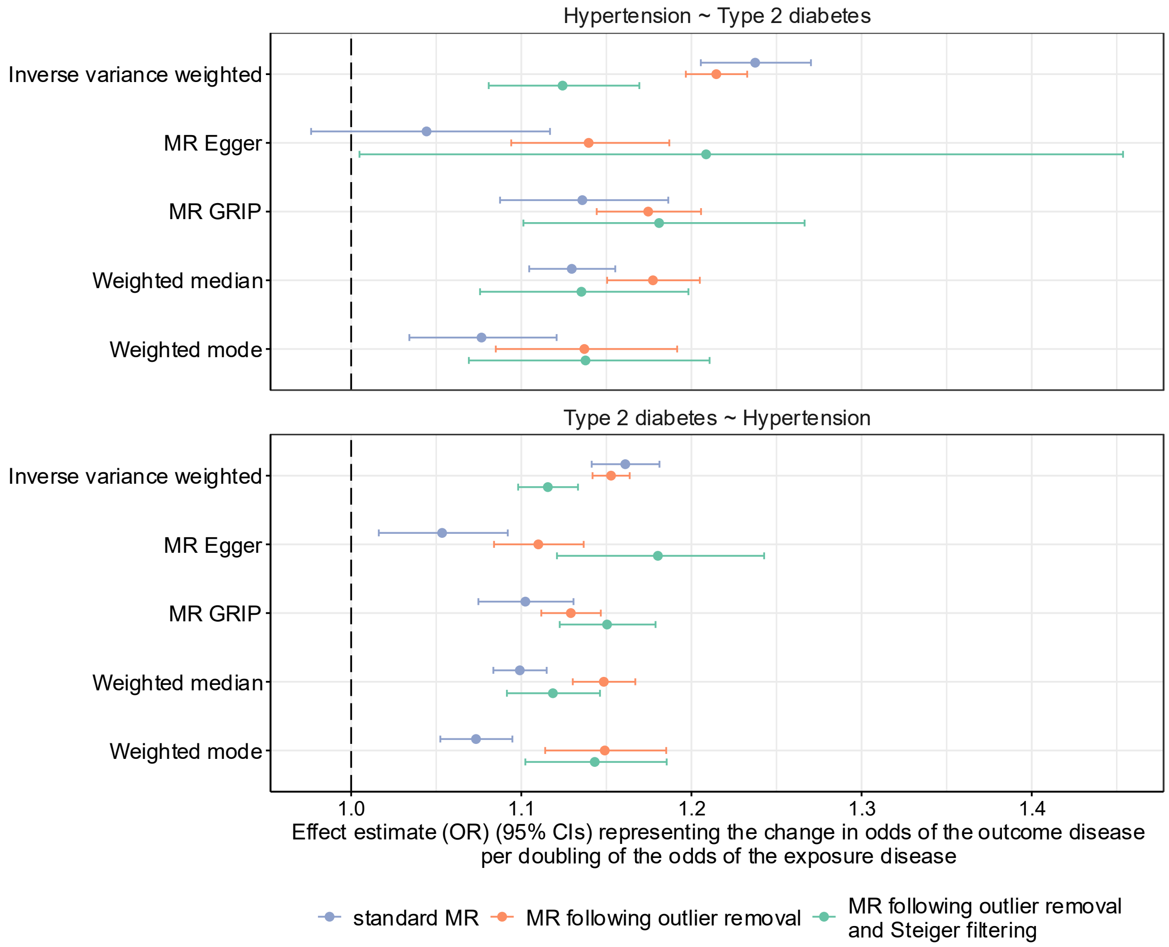

Bidirectional MR estimates for hypertension and T2D. The effect estimates denote the change in odds of the outcome per doubling of the odds of the exposure. MR effect estimates are shown before outlier removal (purple), after outlier removal (orange), and after using Steiger filtering to identify and remove variants that do not explain more variance in the exposure than the outcome (green).

*Supplementary Figure 4: Genetic variant rs10090444 near the gene PINX1, associations with known traits and protein levels*

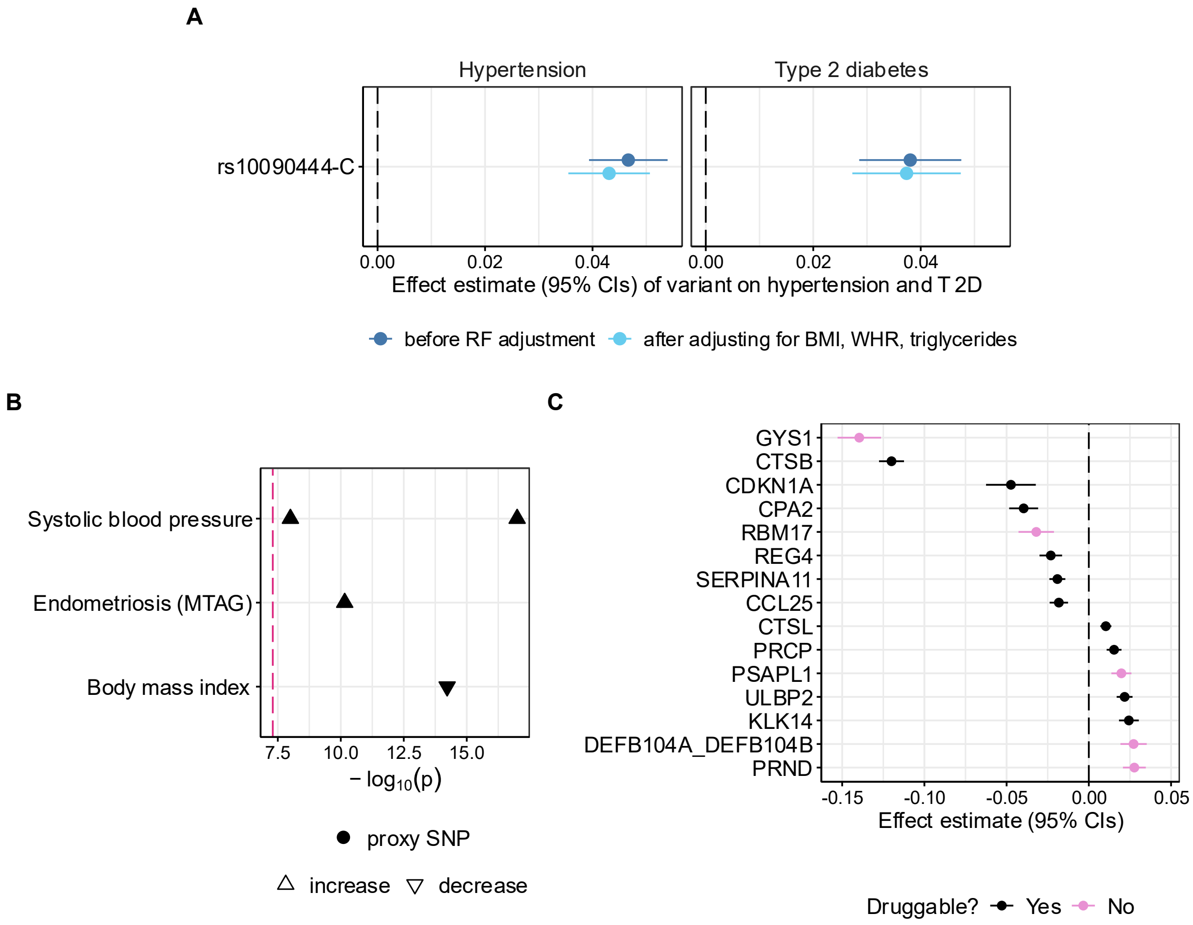

A) rs10090444 associations with hypertension and type 2 diabetes in GEMINI data before and after removing the causal effect of BMI, WHR and triglycerides. B) rs10090444 associations with published results from GWAS Catalog[2] (or a proxy variant in LD with $R^2$>0.9), see Supplementary Table 7 for full results. C) top 15 most significantly associated proteins with rs10090444 in UK Biobank proteomics data[3], see Supplementary Table 8 for full results.

*Supplementary Figure 5: Genetic variant rs5215 in the gene KCNJ11, associations with known traits and protein levels*

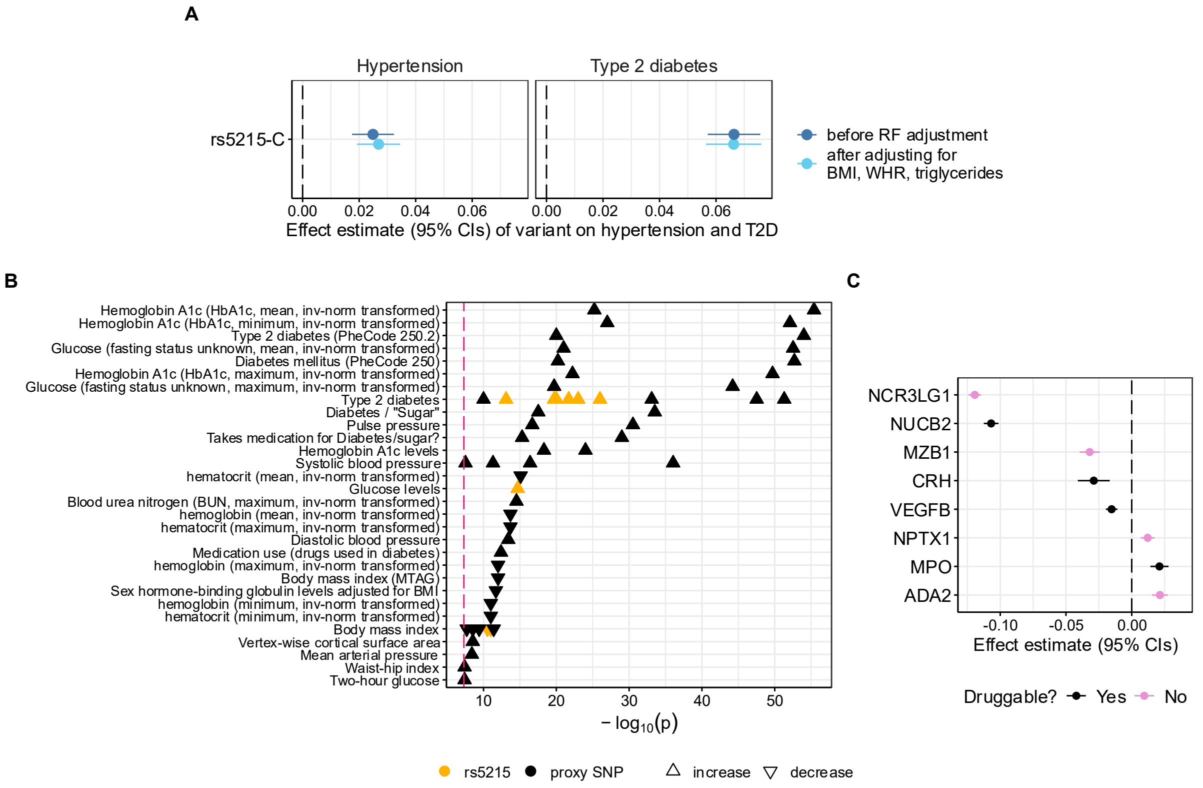

A) rs5215 associations with hypertension and type 2 diabetes in GEMINI data before and after removing the causal effect of BMI, WHR and triglycerides. B) rs5215 associations with published results from GWAS Catalog[2] (or a proxy variant in LD with $R^2$>0.9), see Supplementary Table 7 for full results. C) top 15 most significantly associated proteins with rs5215 in UK Biobank proteomics data[3], see Supplementary Table 8 for full results.

*Supplementary Figure 6: Genetic variant rs56408111 in the gene ZNF101, associations with known traits and protein levels*

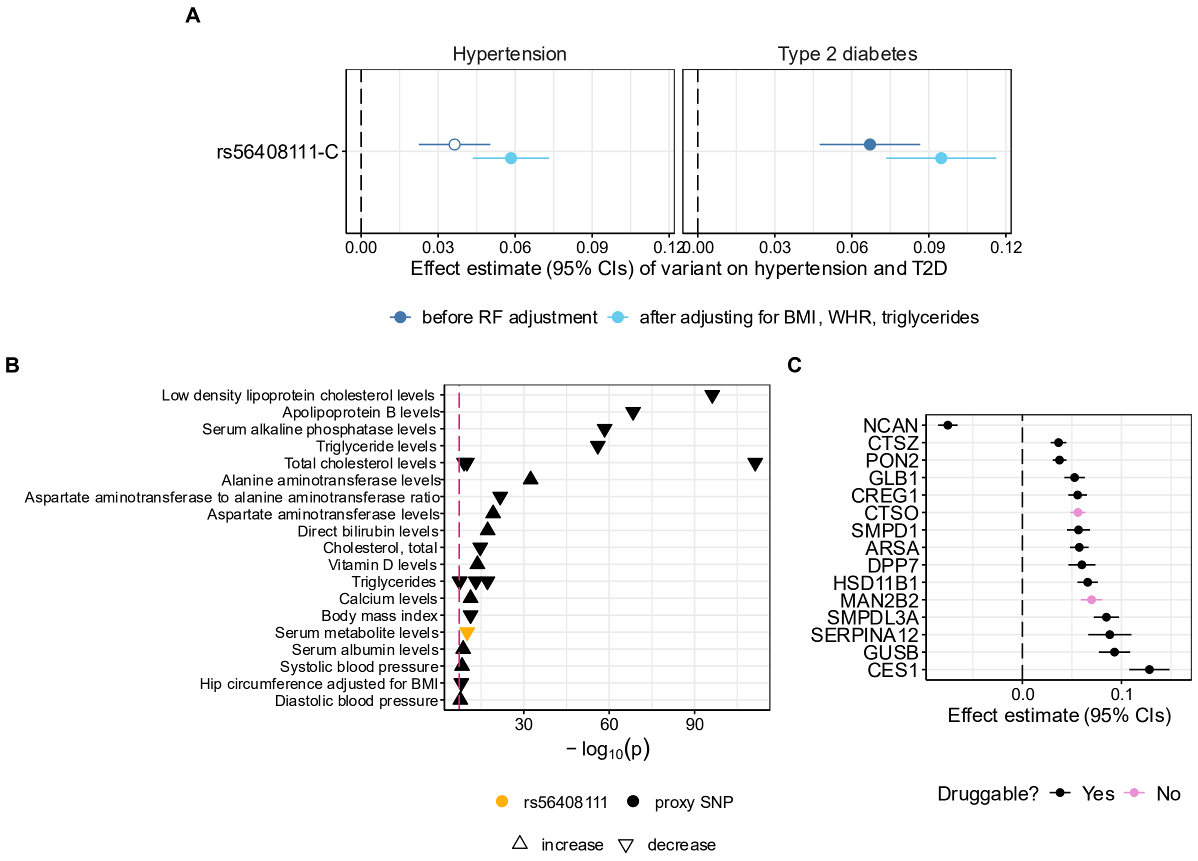

A) rs56408111 associations with hypertension and type 2 diabetes in GEMINI data before and after removing the causal effect of BMI, WHR and triglycerides. B) rs56408111 associations with published results from GWAS Catalog[2] (or a proxy variant in LD with $R^2$>0.9), see Supplementary Table 7 for full results. C) top 15 most significantly associated proteins with rs56408111 in UK Biobank proteomics data[3], see Supplementary Table 8 for full results.

#### Supplementary Methods

*Risk factor GWAS sources*

- BMI[4]
- WHR[5]
- Cholesterol:
    - LDL[6]
    - HDL[6]
    - Triglycerides[6]
- Educational attainment[7]
- Smoking status:
    - Ever smoked[8]
    - Smoking intensity in smokers[8]
- Alcohol consumption[8]
- Physical activity[9]

*Partial LDSC: extension to multiple confounders*

In our previous work, we introduced our method to investigate the role of risk factors, such as BMI, in explaining shared genetics amongst long-term conditions; this was limited to the inclusion of single risk factors at a time [10]. Here, we extend our method to enable the inclusion of multiple confounders, allowing us to quantify the contribution of a set of potential confounders to the genetic similarity between pairs of conditions.

The partial genetic covariance between conditions $k$ and $l$ ($\hat{\rho}_{g\{k,l\,|\,x\}}$), which corresponds to their genetic covariance while holding the genetic effects of a set of $n$ traits, denoted as $x$, constant can be defined as follows:

$$\hat{\rho}_{g\{k,l\,|\,x\}} = \hat{\rho}_{g\{k,l\}} - \hat{\boldsymbol{\rho}}_{g\{k,x\}}\, \hat{\boldsymbol{\rho}}^{-1}_{g\{x,\,x\}} \hat{\boldsymbol{\rho}}_{g\{x,l\}}, \qquad (1)$$

where $\hat{\rho}_{g\{k,l\}}$ is the genetic covariance between condition $l$ and $k$ (1 x 1),

$\hat{\boldsymbol{\rho}}_{g\{k,x\}}$ is the genetic covariance between condition $k$ and traits from $x$ (1 x $n$),

$\hat{\boldsymbol{\rho}}_{g\{x,l\}}$ is the genetic covariance between traits from $x$ and condition $l$ ($n$ x 1),

$\hat{\boldsymbol{\rho}}_{g\{x,\,x\}}$is the matrix of pairwise genetic covariance for all traits from $x$ ($n$ x n).

We have implemented this method in {partialLDSC} R package v0.2.0 (https://github.com/GEMINI-multimorbidity/partialLDSC).

### Supplementary Results

*Estimated causal effect of hypertension on T2D – sensitivity analyses*

Cochrane's Q-statistic was much greater than recommended, indicating that we may be including SNPs that violate the exclusion-restriction assumption, biasing the causal effect estimate. More pleiotropy-robust methods, the weighted median and weighted mode estimators, gave smaller effect estimates, though still statistically significant (Supplementary Table 2). Radial MR identified 163 (out of 547) SNPs as outliers. After removing these SNPs, the IVW estimate reduced slightly: logOR = 0.28; (95% CI: 0.26-0.30; p = $2.23*10^{-145}$), and the Cochrane's Q-statistic reduced from Q = 3377 (546 degrees of freedom [DF]), to Q = 452 (383 DF).

*Estimated causal effect of T2D on hypertension – sensitivity analyses*

Cochrane's Q-statistic again showed evidence of heterogeneity: Q = 3034 (454 DF). After removing 161 (out of 455) outlying SNPs identified by Radial MR, the IVW estimate remained largely unchanged: OR = 1.15 (95% CI: 1.14-1.16; p = 2.26*10-190), with a substantial reduction in the Q-statistic: Q = 303 (293 DF). The individual SNP effects are shown in Supplementary Figure 3, and the sensitivity analysis MR estimates are shown in Supplementary Figure 4.
